# Unquantifiably low aldosterone concentrations are prevalent in hospitalised COVID-19 patients but may not be revealed by chemiluminescent immunoassay

**DOI:** 10.1101/2022.02.28.22271645

**Authors:** Martin Wiegand, David J. Halsall, Sarah L. Cowan, Kevin Taylor, Robert J. B. Goudie, Jacobus Preller, Mark Gurnell

## Abstract

**Objective:** Previous studies have reported conflicting findings regarding aldosterone levels in patients hospitalised with COVID-19. We therefore used the gold-standard technique of liquid chromatography tandem mass-spectrometry (LCMSMS) to address this uncertainty.

**Design:** All patients admitted to Cambridge University Hospitals with COVID-19 between March 10, 2020 and May 13, 2021, and in whom a stored blood sample was available for analysis, were eligible for inclusion.

**Methods:** Aldosterone was measured by LCMSMS and by immunoassay; cortisol and renin were determined by immunoassay.

**Results:** Using LCMSMS, aldosterone was below the limit of detection (<70 pmol/L) in 74 (58.7%) patients. Importantly, this finding was discordant with results obtained using a commonly employed clinical immunoassay (Liaison Diasorin®), which over-estimated aldosterone compared to the LCMSMS assay (intercept 14.1 [95% CI -34.4 to 54.1] + slope 3.16 [95% CI 2.09 to 4.15] pmol/L). The magnitude of this discrepancy did not clearly correlate with markers of kidney or liver function. Solvent extraction prior to immunoassay improved the agreement between methods (intercept -14.9 [95% CI -31.9 to -4.3] and slope 1.0 [95% CI 0.89 to 1.02] pmol/L) suggesting the presence of a water-soluble metabolite causing interference in the direct immunoassay. We also replicated a previous finding that blood cortisol concentrations were often increased, with increased mortality in the group with serum cortisol levels >744 nmol/L (p=0.005).

**Conclusion:** When measured by LCMSMS, aldosterone was found to be profoundly low in a significant proportion of patients with COVID-19 at the time of hospital admission. This has likely not been detected previously due to high levels of interference with immunoassays in patients with COVID-19, and this merits further prospective investigation.

## Introduction

As of February 2022 the United Kingdom has recorded in excess of 18.8 million cases of COVID-19, with at least 161,000 cases resulting in death [1]. The speed of onset and severity of the pandemic has spurred a coordinated response from the global biomedical community on a scale not previously seen. This includes attempts to better understand the pathogenesis of the disease, to identify factors that can be used to predict risk and disease trajectory in individual patients, and to deliver preventative and curative interventions for the world’s population.

Given the key role of angiotensin-converting enzyme 2 (ACE2) in facilitating entry of SARS-CoV-2 virus particles into the lung (alveolar epithelial type II cells), gastrointestinal tract (luminal intestinal epithelial cells) and other tissues [2, 3], exploration of the potential effects on the renin-angiotensin-aldosterone system (RAAS) is of interest in understanding the pathogenesis of COVID-19. ACE2 inhibits RAAS activation by converting angiotensin II (AngII), to angiotensin 1–7 (Ang 1–7). Ang 1–7 exerts anti-inflammatory, anti-oxidative and vasodilatory effects via binding to the Mas receptor [4]. AngII binds AngII receptor type 1 which then exerts pro-inflammatory, pro-oxidative and vasoconstrictive effects [5]. It may also contribute to pro-fibrotic effects, hypercoagulability and immunothrombosis by inducing tissue factor and plasminogen activator inhibitor-1 expression by endothelial cells. AngII further binds to the angiotensin I receptor on the adrenal glands, stimulating the release of the mineralocorticoid aldosterone.

SARS-CoV-2 has the potential to activate RAAS and the secretion of aldosterone, by preventing this ACE2-Ang1–7 mediated RAAS inhibition. The uninhibited Ang II may then play a role in the pathogenesis of the observed hypertension [6], inflammation, immunothrombosis and possible fibrosis in COVID-19.

While elevated serum cortisol has been identified as a marker of poor prognosis in COVID-19 patients [7], the evidence regarding RAAS activation is less clear. Early studies found evidence of increased RAAS activation [8, 9, 10], but subsequent reports suggested no association [11]. Similarly, there is conflicting evidence regarding serum aldosterone levels, with both increased concentrations [12] and no changes reported [11]. In addition, there have been several case reports of hyporeninemic hypoaldosteronism [13]. A low aldosterone/renin ratio has also recently been suggested as predictive of increased severity [14].

To the best of our knowledge, all published studies measuring aldosterone in COVID-19 patients have used non-extraction immunoassays. These methods lack specificity and are prone to interference, for example the polar aldosterone metabolite Aldosterone-18-glucuronide has been shown to cross react in a non-extraction assay commonly used in clinical laboratories [15, 16]. This is more apparent in patients with renal failure as hydrophilic metabolites accumulate. Mass spectrometric methods for aldosterone are now increasingly available in clinical laboratories and do not suffer this interference. Whilst the clinical effectiveness of non-extraction immunoassays in the diagnosis of primary hyperaldosteronism is still contested [17], mass spectrometric methods are metrologically superior and are more likely to represent the biologically active aldosterone fraction.

During the pandemic we observed low aldosterone levels in a number of patients, which we had not anticipated. Motivated by this observation, in this study we used a tandem mass spectrometric method to estimate serum aldosterone concentration in patients admitted to hospital with SARS-CoV-2 infection. We correlate these serum aldosterone results with clinical outcomes, and compare results from the tandem mass spectrometric method with re-measurements using immunoassay methods. We also evaluate the association between high cortisol concentrations and 28-day survival, as previously described by Tan et al [7].

## Methods

All patients presenting to Cambridge University Hospitals, UK, who had a positive diagnostic test for SARS-CoV-2 between March 10, 2020 and May 13, 2021 were eligible for inclusion. Diagnostic testing for SARS-CoV-2 at the hospital used either a real-time reverse transcription polymerase chain reaction (RT-PCR) of the RdRp gene from a nasopharyngeal swab, or the SAMBA II point-of-care test [18].

We made use of blood samples drawn from a biobank at Cambridge University Hospitals, which was established early in the pandemic to store, where available, blood samples from patients with COVID-19. We retrospectively measured aldosterone, cortisol and renin levels on the available stored samples from the biobank. All patients with at least one stored sample from around the time of their first positive SARS-CoV-2 result (within 72 hours) were included. Within this time window, the earliest available sample was used. Patients were excluded if they had received glucocorticoid or mineralocorticoid therapy (e.g. dexamethasone, fludrocortisone, hydrocortisone, methylprednisolone, prednisolone) prior to collection of the blood sample.

Aldosterone was measured using a liquid chromatography tandem mass spectrometric method (LCMSMS) adapted from [19], with a lower limit of quantitation (LLOQ) of 70 pmol/L. The assay in the clinical laboratory is accredited by UKAS (https://www.ukas.com/) to the ISO 15189 standard. Serum cortisol was measured using Siemens Centaur direct competitive CLIA (LLOQ 25 nmol/L). On a subset of patients where appropriately stored plasma samples were available (samples not chilled prior to centrifugation and plasma stored frozen) we also measured plasma renin concentration, using the Diasorin LIAISON® immunometric CLIA method (LLOQ 1.96 µIU/mL).

The association between aldosterone, cortisol and renin in paired samples was assessed by linear regression, imputing the value 35 pmol/L for aldosterone values below the LLOQ of 70 pmol/L, which is statistically equivalent to assuming that these values are uniformly distributed between 0 and the LLOQ [20].

To assess the association between high cortisol concentrations (> 744 nmol/L) and outcomes [7], a Kaplan-Meier estimate of mortality within 28-days of a positive SARS-CoV-2 test, stratified by the first cortisol concentration following the positive test (within 72 hours), was used. Tan et al. selected the threshold of 744 nmol/L as the value that maximises the log-rank statistic, i.e. the difference between survival curves. The selection of the cortisol threshold that optimises the difference between survival curves in our cohort was also replicated and the difference in survival curves was assessed by a log-rank test. Similarly the difference in survival curves of cohorts with high or low aldosterone concentrations was investigated.

To explore potential differences between aldosterone as determined by LCMSMS vs immunoassay, aldosterone was re-measured in a subset of patients, where sample volume permitted, using the Diasorin LIAISON® direct competitive chemiluminescent immunoassay (CLIA) (LLOQ 1.91 ng/dL). Scatter plots, with Passing-Bablok regression lines, and Bland-Altmann plots were used to assess the agreement between the LCMSMS and CLIA methods. To explore potential association between bias in the CLIA results with renal function, we examined associations with estimated glomerular filtration rate (eGFR) and creatinine clearance (Cockroft-Gault) using linear regression that is robust to outliers [21].

Finally, the CLIA was repeated following solvent extraction of serum to remove potential aqueous interference from patient blood samples that would not be detected by the LCMSMS method. In brief, 350ul of patient sample was mixed with 2250ul of Methyl Tertiary Butyl Ether by vortex for 15 minutes. The lower aqueous layer was frozen and the upper MTBE layer decanted and evaporated to dryness at 60°C under nitrogen flow. The sample was reconstituted in 350ul of steroid free serum (DRG Instruments GmbH Marburg Germany). Extraction efficiency was corrected for using the mean extraction recovery from a cohort of 15 anonymised COVID-19 negative control subjects (data not shown).

## Results

231 patients who tested positive for SARS-CoV-2 had at least one measurement of aldosterone, cortisol or renin within 72 hours of their first positive SARS-CoV-2 test. 97 patients had received glucocorticoid and/or mineralocorticoid therapy prior to the collection of the blood sample, leaving 134 patients in the study cohort (126 with an aldosterone measurement available; 88 with measured cortisol; and 50 with measured renin).

80 patients (59%) were male and the median age was 64 years (IQR 46, 88). 15 patients (11.2%) were admitted to the intensive care unit (ICU) and the in-hospital mortality was 13.4%. Other baseline parameters and markers of severity are described in Table 1. The patients in our study had similar age, gender, ethnicity, BMI and severity markers to the overall population of SARS-CoV-2 patients presenting to the hospital (eTable 1), although these patients had lower interleukin-6 (median 8.1 vs 11.9); were less likely to be admitted to ICU (11.2% vs 18.9%); and were more likely to have diabetes (32.8% vs 20.7%) or other endocrine disease (14.2% vs 8.7%).

**Table 1.** Patient characteristics following their first positive SARS-CoV-2 test.

| Characteristic |  |
| --- | --- |
| Sample size (n) | 134 |
| Age at admission, median [IQR] (range) | 64 [46, 88] (20—102) |
| Gender (male), n (%) | 80 (59%) |
| ***Ethnicity, n (%)*** |  |
| White | 90 (67.2%) |
| Black | 1 (0.7%) |
| Asian | 13 (9.7%) |
| Other | 3 (2.2%) |
| Not specified/prefer not to say | 27 (20.1%) |
| Body mass index, median [IQR], kg/m | 27.8 [23.5, 32.3] |
| ***Observations, median [IQR]*** |  |
| Heart rate, beats/min | 86 [74, 95] |
| Temperature, °C | 37.1 [36.6, 37.7] |
| Respiratory rate, breaths/min | 19 [17, 22] |
| Oxygen saturation (SpO<sub>2</sub>), % | 96 [94, 98] |
| Mean arterial pressure, mmHg | 90 [82, 99] |
| ***Blood tests, median [IQR]*** |  |
| C-reactive protein, mg/L | 51 [19, 107] |
| White cell count, 10<sup>9</sup>/L | 6.2 [5, 8.9] |
| Sodium, mmol/L | 137.8 [135.6, 140.0] |
| Potassium, mmol/L | 4.0 [3.8, 4.5] |
| Neutrophils, 10<sup>9</sup>/L | 4.7 [3.4, 7.0] |
| Lymphocytes, 10<sup>9</sup>/L | 1.0 [0.7, 1.5] |
| Interleukin-6, pg/ml | 8.1 [3.6, 18.5] |
| Urea, mmol/L | 5.8 [4.2, 8.8] |
| Creatinine, μmol/L | 72 [62, 93] |
| D-Dimer, ng/ml | 221 [142, 461] |
| Troponin, ng/L | 8.5 [3.0, 24.7] |
| pH value | 7.40 [7.37, 7.44] |
| ***Medical history, n (%)*<sup>1</sup>** |  |
| Heart disease | 24 (17.9%) |
| Hypertension | 48 (35.8%) |
| Beta blockers at admission<sup>2</sup> | 27 (20.2%) |
| Diabetes | 44 (32.8%) |
| Endocrine disease (other than diabetes) | 19 (14.2%) |
| Stroke | 3 (2.2%) |
| Dementia | 10 (7.5%) |
| Asthma | 20 (14.9%) |
| Respiratory disease (other than asthma) | 17 (12.7%) |
| Chronic kidney disease | 9 (6.7%) |
| Chronic liver disease | 10 (7.5%) |
| Malignancy non-haematological | 18 (13.4%) |
| Malignancy haematological | 4 (3.0%) |
| Immunocompromised | 2 (1.5%) |
| ***Treatments and outcomes, n (%)*** |  |
| In-hospital deaths | 18 (13.4%) |
| Admitted to ICU | 15 (11.2%) |
| Invasive mechanical ventilation | 12 (8.9%) |
| Renal replacement therapy | 10 (7.5%) |

<sup>1</sup>See eTable 3 for ICD-10 code lists.
<sup>2</sup>Bisoprolol, Atenolol, Propranolol, Carvedilol.

The distribution of aldosterone, cortisol and renin is visualised by histograms in Figure 1. The aldosterone concentrations after the positive test in Figure 1A are remarkably low, with the concentration of aldosterone below the LLOQ (70 pmol/L) in 58.7% of the patients (74 patients). Analysis of cortisol levels found elevated levels, with 20.5% of patients having results greater than 744 nmol/L, the level proposed by Tan el al to maximise the difference between patient survival curves (Figure 1B). Figure 1C depicts the renin concentrations, which were predominantly low despite the low aldosterone concentration.

**Figure 1.**
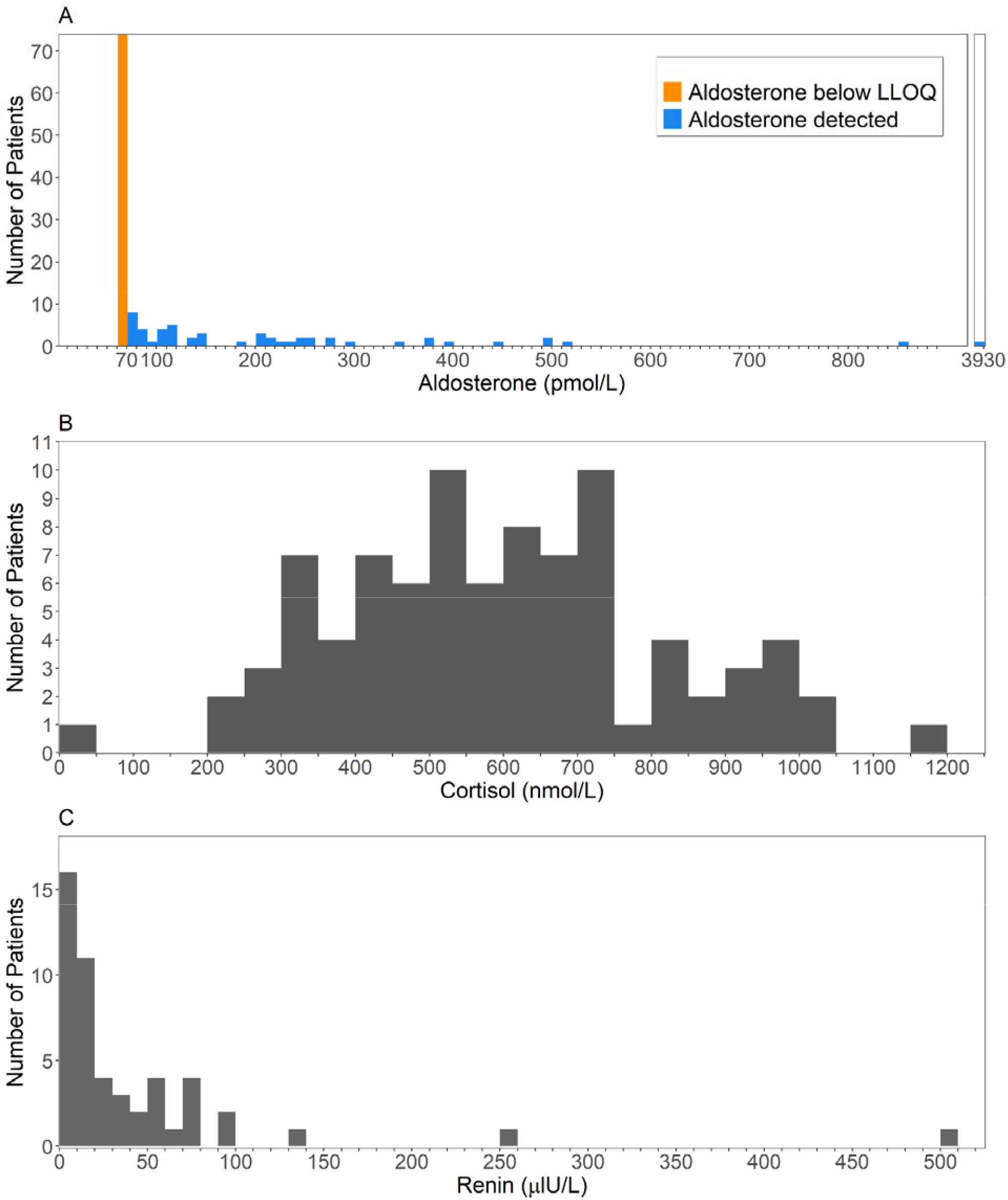
Histograms of the first available result (**A)** Aldosterone, measured using LCMSMS (**B)** Cortisol (**C)** Renin.

There was evidence that higher aldosterone concentrations were associated with both higher cortisol (beta=0.316, 95% CI 0.06 to 0.57, p=0.02) and higher renin concentrations (beta=0.062, 95% CI 0.025 to 0.099, p<0.00001), as shown in Figure 2. Stratification of the cohort according to whether aldosterone was above or below the LLOQ did not reveal any significant differences in clinical characteristics (eTable 2), or in the survival curves (eFigure 2). There was no obvious correlation between aldosterone level and renal function (eFigure 3).

**Figure 2.**
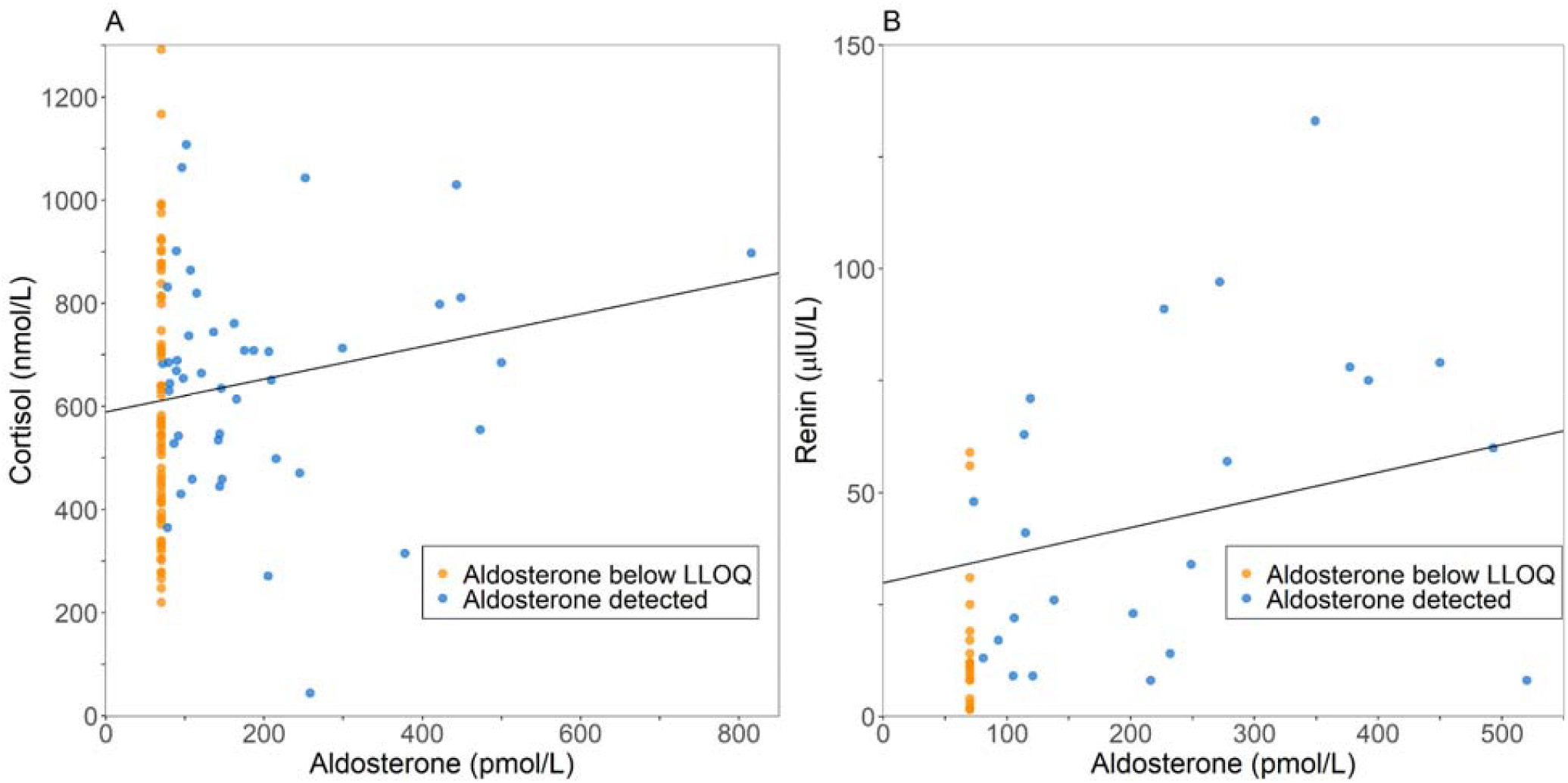
Scatter plots of paired aldosterone, cortisol and renin results, with linear regression lines shown. (A) Aldosterone (measured using LCMSMS) against cortisol. (B) Aldosterone (LCMSMS) against renin (two extreme outliers are outside the plotting region; see eFigure 1 for zoomed-out version).

The survival probability of patients with a high concentration of cortisol (above 744 nmol/L) was lower than for those with a low concentration of cortisol (below 744 nmol/L). As shown in Figure 3, there was a marked difference in 28-day mortality, with 44% of patients dying in the high cortisol group, compared to 11% of patients in the low cortisol group. While the 95% confidence intervals (CI) around the survival curves overlap over the entire time frame, the log-rank test comparison of the complete survival curves indicates a significant difference between the two groups (p = 0.005). The average length of hospital stay in the high cortisol group (19.1 days) was longer than in the low cortisol group (11.2 days). The cortisol threshold maximising the log-rank test for the difference of survival curves in our cohort was 801 nmol/L.

**Figure 3.**
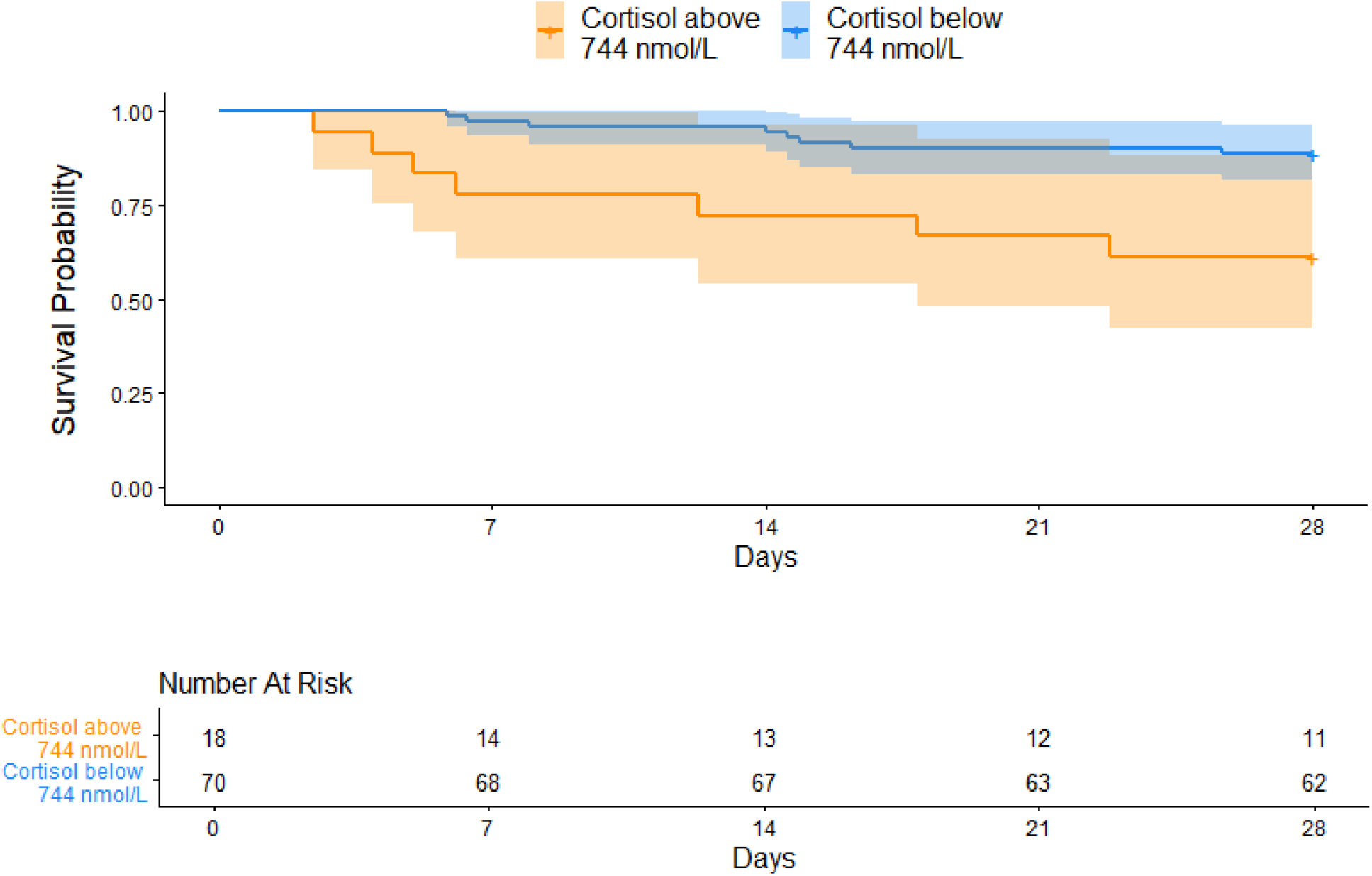
Kaplan-Meier survival plot for 28-day survival in steroid naive patients after first positive test, stratified by the first cortisol result after the test. The 95% confidence intervals are shown by shading.

The considerable disparity between our LCMSMS aldosterone results and previously-published studies [12, 11], which used immunoassay, is explained by Figure 4. This shows that re-measurements of aldosterone by CLIA displayed significant proportional positive bias compared to the LCMSMS results: CLIA (pmol/L) = 14.1 + 3.16 × LCMSMS (pmol/L), with intercept 95% CI -34.4 to 54.1 and slope 95% CI 2.09 to 4.15. The Bland-Altmann plot (eFigure 4) further highlights the large mean difference between the methods (319.5 pmol/L), wide limits of agreement, and suggests the differences increase as the average of the LCMSMS and CLIA measurements increases. No clear explanation for the difference between LCMSMS and CLIA results were observed in markers of renal or liver function (eFigure 5).

**Figure 4.**
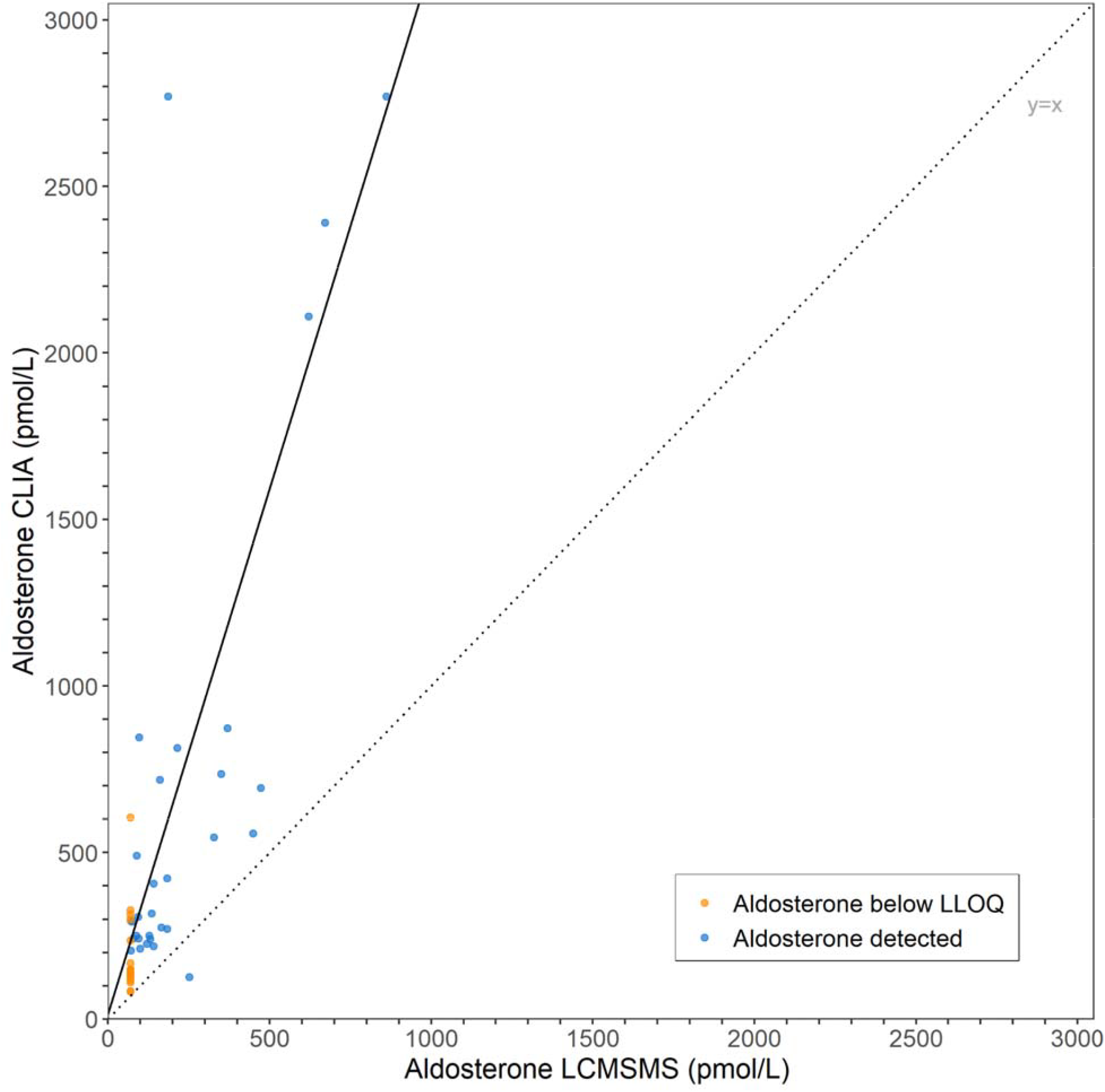
Comparison of aldosterone measured by LCMSMS and CLIA with Passing-Bablok regression line (solid black) shown. The dotted gray line indicates the y=x identity line.

The agreement between methods was considerably improved following solvent extraction of serum, as shown in eFigure 6: Extracted CLIA = -14.9 + 1.0 × LCMSMS (intercept 95% CI -31.9 to -4.3 and slope 95% CI 0.9 to 1.0), with reduced random noise (Pearson R^2^ = 0.97 cf. 0.60), suggesting interference in the immunoassay by a water-soluble metabolite.

## Discussion

Given the role of ACE2 and the renin-angiotensin-aldosterone system (RAAS) in COVID-19 [6], the finding of low aldosterone concentration is surprising and in conflict with previous studies that have deployed immunoassays to measure aldosterone in COVID-19 patients. Whilst there are case reports of hyporeninemic hypoaldosteronism in COVID-19 [13] these patients do not generally present with clinical features of hypoaldosteronism, suggesting that despite the lack of correlation between serum aldosterone and cortisol concentration, sufficient mineralocorticoid activity is present to avoid decompensation. Other mechanisms to consider include activation of epithelial sodium channels (ENaC) in the distal nephrons by AngII as opposed to aldosterone, increasing sodium reabsorption and volume expansion (at the cost of potassium and hydrogen ion excretion), which may then provide further negative feedback for RAAS, with associated hyporeninemia and hypoaldosteronism while AngII over-expression continues to be facilitated by ACE2 inhibition.

Our data do reproduce the findings of Tan et al [7], that serum cortisol is elevated in COVID-19 patients and is a negative prognostic indicator. In addition to activating the glucocorticoid receptor (GR), cortisol is also an effective ligand for the mineralocorticoid receptor (MR). In normal physiology 11β-hydroxysteroid dehydrogenase type 2 (11β-HSD2) inactivates cortisol to cortisone thus preventing cross talk at this receptor. However, when cortisol levels are significantly raised, 11β-HSD2 is saturated and activation of MR ensues. Accordingly, a possible explanation of the counter-intuitive observation of low serum aldosterone concentration without clinically apparent hypoaldosteronism is that the high serum cortisol concentrations found in these patients has sufficient mineralocorticoid activity to suppress RAAS. However, the weak positive correlation between aldosterone and cortisol in these patients does not fully support this finding and a more complex explanation is required. An alternative explanation could be that the polar aldosterone metabolites have activity at the mineralocorticoid receptor as well as cross reacting in the non-extraction immunoassay. However, previous studies suggest that if aldosterone metabolites do bind to the mineralocorticoid receptor, they do so with much reduced affinity [22].

The high prevalence of undetectably-low aldosterone concentration in patients with COVID-19 has not been previously reported, but prior studies have reported results from immunoassay rather than mass spectroscopy [23, 14]. The limited agreement in aldosterone results between CLIA and LCSMS is evident. Whilst this poor correlation has been previously established [24, 25], the effect in COVID-19 patients is exaggerated. Solvent extraction significantly improves the correlation between methods suggesting a water soluble metabolite may be cross reacting in the direct immunoassay; this has also been previously demonstrated using extraction immunoassay [15]. Aldosterone-18-glucuronide [17], the principal metabolite of aldosterone, is a likely candidate. Increases in this metabolite are unlikely to be a direct effect of COVID-19 infection, but may reflect secondary organ dysfunction due to systemic illness. Whilst renal impairment has been shown indirectly to increase aldosterone glucuronide [26], a correlation between eGFR and the discrepancy between the two assays was not demonstrable in this cohort.

To our knowledge, this study is the first to report aldosterone levels in COVID-19 patients as measured by LCMSMS, along with re-measurements using CLIA. A weakness of our study is that, whilst it appears that our sample is representative of the broader cohort of hospitalised COVID-19 patients in terms of clinical characteristics at admission, our sample is a convenience sample of patients with suitable available samples in the hospital’s biobank, rather than a prospectively-collected cohort. This also meant we were unable to measure renin in all patients, due to the more specific sample requirements. Nonetheless, we believe our findings to be important and hope they will spur further research on the role of RAAS in COVID-19.

In conclusion, these data demonstrate that aldosterone cannot be accurately estimated in serum from patients with SARS-CoV-2 infection using direct competitive immunoassay due to the presence of a water soluble interference. When measured using gold-standard LCMSMS, serum aldosterone is found to be remarkably low in most patients with COVID-19. The mechanism of this reduction remains obscure with no obvious correlation with glucocorticoid status or kidney function.

## Data Availability

The de-identified data that support the findings of this study are available from Cambridge
University Hospitals but restrictions apply to the availability of these data, which were used
under license for the current study, and so are not publicly available. Data are however
available from the authors upon reasonable request with permission of Cambridge University
Hospitals.

## Acknowledgements

We are grateful to Professor Christopher Edwards, Professor Morris Brown and Professor Will Drake for helpful discussions in the initial phase of this work. The clinical informatics data extraction was funded by the Cancer Research UK Cambridge Center and conducted by Vince Taylor.

Martin Wiegand was funded by the NIHR Cambridge Biomedical Research Centre [BRC-1215-20014]. Robert J. B. Goudie was funded by the UKRI Medical Research Council (MRC) [programme code MC_UU_00002/2] and supported by the NIHR Cambridge Biomedical Research Centre [BRC-1215-20014]. Mark Gurnell was supported by the NIHR Cambridge Biomedical Research Centre [BRC-1215-20014].

The funders had no role in the design and conduct of the study; collection, management, analysis, and interpretation of the data; preparation, review, or approval of the manuscript; and decision to submit the manuscript for publication. The views expressed are those of the authors and not necessarily those of the NHS, the NIHR, or the MRC.

## Author contributions

**Conception:** MG, DJH and JP conceived the research project and designed the study.

**Data collection:** DJH, MW, SLC and RJBG extracted and curated the dataset. DJH and KT provided the laboratory analysis.

**Analysis tools & Data interpretation:** MW, RJBG and analysed and contributed to the selection and creation of analysis tools. MG, JP, SLC and DJH interpreted the data and results.

**Implementation:** MW, RJBG and DJH performed the implementation and analysis of the proposed method.

**Draft writing:** DJH, MW, SLC and RJBG wrote the initial draft. All authors contributed to substantively revising the article for important intellectual content.

### Approval of final submission

All authors approved of the final version of this script to be published, and are accountable for the work presented.

## References

[1] http://Gov.Uk dashboard to track COVID-19 cases in the United Kingdom. 28/02/2022. https://coronavirus.data.gov.uk/details/deaths?areaType=overview&areaName=United%20Kingdom

[2] Zhou P, Yang X-L, Wang X-G, Hu B, Zhang L & Zhang W. A pneumonia outbreak associated with a new coronavirus of probable bat origin. Nature. 2020 579 270–273. https://doi.org/10.1038/s41586-020-2012-7

[3] Hoffmann M, Kleine-Weber H, Schroeder S, Krüger N, Herrler T & Erichsen S. SARS-CoV-2 cell entry depends on ACE2 and TMPRSS2 and is blocked by a clinically proven protease inhibitor. Cell. 2020 181(2) 271–280.e8. https://doi.org/10.1016/j.cell.2020.02.052

[4] Santos RA, Ferreira AJ, Verano-Braga T & Bader M. Angiotensin-converting enzyme 2, angiotensin-(1-7) and Mas: new players of the renin-angiotensin system. Journal of Endocrinology. 2013 216(2) R1–R17. https://doi.org/10.1530/JOE-12-0341

[5] Vaduganathan M, Vardeny O, Michel T, McMurray JJV, Pfeffer MA & Solomon SD. Renin-Angiotensin-Aldosterone System Inhibitors in Patients with Covid-19. New England Journal of Medicine. 2020 382(17) 1653–1659. https://doi.org/10.1056/NEJMsr2005760

[6] Rysz S, Al-Saadi J, Sjöström A, Farm M, Jalde FC & Plattén M. COVID-19 pathophysiology may be driven by an imbalance in the renin-angiotensin-aldosterone system. Nature Communications. 2021 12 2417. https://doi.org/10.1038/s41467-021-22713-z

[7] Tan T, Khoo B, Mills EG, Phylactou M, Patel B & Eng PC. Association between high serum total cortisol concentrations and mortality from COVID-19. The Lancet Diabetes & Endocrinology. 2020 8(8) 659–660. https://doi.org/10.1016/S2213-8587(20)30216-3

[8] Wu Z, Hu R, Zhang C, Ren W, Yu A & Zhou X. Elevation of plasma angiotensin II level is a potential pathogenesis for the critically ill COVID-19 patients. Critical Care. 2020 24 290. https://doi.org/10.1186/s13054-020-03015-0

[9] Liu N, Hong Y, Chen R-G & Zhu H-M. High rate of increased level of plasma Angiotensin II and its gender difference in COVID-19: an analysis of 55 hospitalized patients with COVID-19 in a single hospital, WuHan, China. medRxiv. 2020.04.27.20080432; https://doi.org/10.1101/2020.04.27.20080432

[10] Dudoignon E, Moreno N, Deniau B, Coutrot M, Longer R & Amiot Q. Activation of the renin-angiotensin-aldosterone system is associated with Acute Kidney Injury in COVID-19. Anaesthesia Critical Care & Pain Medicine. 2020 39(4) 453–455. https://doi.org/10.1016/j.accpm.2020.06.006

[11] Henry BM, Benoit S, Lippi G, Benoit J. Circulating plasma levels of angiotensin II and aldosterone in patients with coronavirus disease 2019 (COVID-19): A preliminary report. Progress in Cardiovascular Diseases. 2020 63(5) 702–703. https://doi.org/10.1016/j.pcad.2020.07.006

[12] Villard O, Morquin D, Molinari N et al. The plasmatic aldosterone and c-reactive protein levels, and the severity of Covid-19: The Dyphor-19 study. 2020. Journal of Clinical Medicine 9 (7): 2315.

[13] Mandal AKJ, Kho J, Metaxa S & Missouris CG. COVID-19 and late-onset hypertension with hyporeninaemic hypoaldosteronism. The International Journal of Clinical Practice. 2020 75(1) e137732020. https://doi.org/10.1111/ijcp.13773

[14] Akin S, Schriek P, van Nieuwkoop C & Neuman RI. A low aldosterone/renin ratio and high soluble ACE2 associate with COVID-19 severity. Journal of Hypertension. 2022 40(3) 606–614 https://doi.org/10.1097/HJH.0000000000003054

[15] Jones JC, Cater GD & MacGregor GA. Interference by polar metabolites in a direct radioimmunoassay for plasma aldosterone. Annals of Clinical Biochemistry. 1981 18(1) 54–59. https://doi.org/10.1177/000456328101800111

[16] Lam L, Chiu WW & Davidson JS. Overestimation of aldosterone by immunoassay in renal impairment. Clinical Chemistry. 2016 62(6) 890–891. https://doi.org/10.1373/clinchem.2016.255737

[17] Blocki F, Zierold C, Olson G, Seeman J, Cummings S & Bonelli F. In defense of aldosterone measurement by immunoassay: a broader perspective. Clinical Chemistry and Laboratory Medicine. 2017 55(4) e87–e89. https://doi.org/10.1515/cclm-2016-0707

[18] Assennato SM, Ritchie AV, Nadala C, Goel N, Tie C & Nadalaet LM. Performance evaluation of the SAMBA II SARS-CoV-2 test for point-of-care detection of SARS-CoV-2. Journal of Clinical Microbiology. 2020 59(1) e01262–20. https://doi.org/10.1128/JCM.01262-20

[19] Hinchliffe E, Carter S, Owen LJ & Keevil BG. Quantitation of aldosterone in human plasma by ultra high performance liquid chromatography tandem mass spectrometry. Journal of Chromatography B. 2013 913-914 19–23. https://doi.org/10.1016/j.jchromb.2012.11.013

[20] Richardson DB & Ciampi A. Effects of exposure measurement error when an exposure variable is constrained by a lower limit. American Journal of Epidemiology. 2003 157(4) 355–363. https://doi.org/10.1093/aje/kwf217rgg

[21] PJ Huber. Robust Statistics, John Wiley & Sons. New York, USA, 1981.

[22] Morris DJ, Kenyon CJ, Latif SA, McDermott M, Goodfriend TL. The possible biological role of aldosterone metabolites. Hypertension. 1983 5 (2 Pt 2) https://doi.org/10.1161/01.hyp.5.2_pt_2.i35

[23] Rieder M, Wirth L, Pollmeier L, Jeserich M, Goller I & Baldus N. Serum ACE2, angiotensin II, and aldosterone concentrations are unchanged in patients with COVID-19. American Journal of Hypertension. 2021 34(3) 278–281. https://doi.org/10.1093/ajh/hpaa169

[24] Ray JA, Kushnir MM, Palmer J, Sadjadi S, Rockwood AL & Meikle AW. Enhancement of specificity of aldosterone measurement in human serum and plasma using 2D-LC–MS/MS and comparison with commercial immunoassays. Journal of Chromatography B. 2014 970 102–107. https://doi.org/10.1016/j.jchromb.2014.08.042

[25] Debeljak Z, Marković I, Šerić V, Horvat V, Mandić S & Mandić S. Analytical bias of automated immunoassays for six serum steroid hormones assessed by LC-MS/MS. Biochemia Medica (Zagreb). 2020 30(3) 030701. https://doi.org/10.11613/BM.2020.030701

[26] Koshida H, Miyamori I, Miyazaki R, Tofuku Y & Takeda R. Falsely elevated plasma aldosterone concentration by direct radioimmunoassay in chronic renal failure. The Journal of Laboratory and Clinical Medicine. 1989 114(3) 294–300.

