## Supplemental material for "Unquantifiably low aldosterone concentrations are prevalent in hospitalised COVID-19 patients but may not be revealed by chemiluminescent immunoassay"

### Supplement

#### eTable 1. Patient characteristics after first positive SARS-CoV-2 test.

| **Characteristic** | **All SARS-CoV-2 positive patients** | **Our cohort** | **SARS-CoV-2 positive patients not in our cohort** | **p-value^1^** |
| --- | --- | --- | --- | --- |
| Sample size (*n*) | 1801 | 134 | 1667 | - |
| Age at admission, median [IQR] (% missing) | 65 [49, 79] (0%) | 64 [46, 88] (0%) | 66 [49, 79] (0%) | 0.24 |
| Gender (male), *n* (%) | 973 (54.0%) | 80 (59%) | 893 (53.6%) | 0.20 |
| ***Ethnicity, n (%)*** |  |  |  |  |
| White | 1254 (69.6%) | 90 (67.2%) | 1164 (69.8%) | 0.58 |
| Black | 23 (1.3%) | 1 (0.7%) | 22 (1.3%) | 0.87 |
| Asian | 95 (5.3%) | 13 (9.7%) | 82 (4.9%) | 0.029 |
| Other | 61 (3.4%) | 3 (2.2%) | 58 (3.5%) | 0.61 |
| Not specified/prefer not to say | 368 (20.4%) | 27 (20.1%) | 341 (20.4%) | ~1 |
| Body mass index, median [IQR] (% missing), kg/m | 27.3 [23.5, 31.7] (19.8%) | 27.8 [23.5, 32.3] (11.9%) | 27.3 [23.6, 31.7] (20.4%) | 0.32 |
| ***Observations, median [IQR] (% missing)*** |  |  |  |  |
| Heart rate, beats/min | 85 [74, 96] (11.5%) | 86 [74, 95] (7.5%) | 88 [80, 99] (11.8%) | 0.38 |
| Temperature, ℃ | 37.1 [36.6, 37.7] (11.5%) | 37.1 [36.6, 37.7] (7.5%) | 37.1 [36.6, 37.8] (11.8%) | 0.68 |
| Respiratory rate, breaths/min | 18 [17, 21] (11.5%) | 19 [17, 22] (7.5%) | 18 [17, 21] (11.8%) | 0.71 |
| Oxygen saturation (SpO2), % | 96 [94, 98] (11.6%) | 96 [94, 98] (7.5%) | 96 [94, 98] (11.9%) | 0.73 |
| Mean arterial pressure, mmHg | 89 [80, 99] (11.5%) | 90 [82, 99] (7.5%) | 88 [81, 100] (11.8%) | 0.93 |
| ***Blood tests, median [IQR] (% missing)*** |  |  |  |  |
| C-reactive protein, mg/L | 43 [14, 101] (1.2%) | 51 [19, 107] (1.5%) | 43 [14, 100] (1.2%) | 0.21 |
| White cell count, 10^9^/L | 6.4 [4.8, 8.9] (0.5%) | 6.2 [5,8.9] (0%) | 6.4 [4.8, 8.9] (0.5%) | 0.29 |
| Sodium, mmol/L | 137.5 [135, 140] (1.1%) | 137.8 [135.6, 140] (0%) | 137.5 [135, 140] (1.2%) | 0.71 |
| Potassium, mmol/L | 4 [3.7, 4.3] (1.5%) | 4 [3.8, 4.5] (1.5%) | 4 [3.7, 4.4] (1.5%) | 0.31 |
| Neutrophils, 10^9^/L | 4.7 [3.3, 7] (0.5%) | 4.7 [3.4, 7] (0%) | 4.7 [3.3, 6.9] (0.5%) | 0.56 |
| Lymphocytes, 10^9^/L | 0.9 [0.6, 1.4] (0.5%) | 1.0 [0.7, 1.5] (0%) | 0.9 [0.6, 1.4] (0.5%) | 0.21 |
| Interleukin-6, pg/ml | 11.2 [3, 30.3] (71.5%) | 8.1 [3.6, 18.5] (64.2%) | 11.9 [3, 32.4] (72%) | 0.011 |
| Urea, mmol/L | 6.2 [4.4, 9.4] (5.1%) | 5.8 [4.2, 8.8] (0.7%) | 6.2 [4.5, 9.4] (5.5%) | 0.27 |
| Creatinine, 𝜇mol/L | 72 [58.5, 93] (0.8%) | 72 [62, 93] (0.7%) | 72 [58, 93] (0.8%) | 0.11 |
| D-Dimer, ng/ml | 268 [145, 590] (41.1%) | 221 [142, 461] (38.8%) | 272 [145, 595] (41.2%) | 0.62 |
| Troponin, ng/L | 10 [3, 33] (36.9%) | 8.5 [3, 24.7] (31.3%) | 10.3 [3, 34] (37.3%) | 0.82 |
| pH value | 7.40 [7.37, 7.43] (6.7%) | 7.40 [7.37, 7.44] (3.7%) | 7.40 [7.37, 7.43] (7%) | 0.53 |
| *Past medical history*^2^*, n (%)* |  |  |  |  |
| Heart disease | 347 (19.3%) | 24 (17.9%) | 323 (19.4%) | 0.76 |
| Hypertension | 656 (36.4%) | 48 (35.8%) | 608 (36.5%) | 0.95 |
| Diabetes | 389 (21.6%) | 44 (32.8%) | 345 (20.7%) | 0.0015 |
| Endocrine disease (other than diabetes) | 164 (9.1%) | 19 (14.2%) | 145 (8.7%) | 0.05 |
| Stroke | 68 (3.8%) | 3 (2.2%) | 65 (3.9%) | 0.46 |
| Dementia | 151 (8.4%) | 10 (7.5%) | 141 (8.5%) | 0.81 |
| Asthma | 244 (13.5%) | 20 (14.9%) | 224 (13.4%) | 0.72 |
| Respiratory disease (other than asthma) | 218 (12.1%) | 17 (12.7%) | 201 (12.1%) | 0.94 |
| Chronic kidney disease | 141 (7.8%) | 9 (6.7%) | 132 (7.9%) | 0.74 |
| Chronic liver disease | 96 (5.4%) | 10 (7.5%) | 86 (5.2%) | 0.35 |
| Malignancy non-haematological | 246 (13.7%) | 18 (13.4%) | 228 (13.7%) | ~1 |
| Malignancy haematological | 87 (4.8%) | 4 (3.0%) | 83 (5.0%) | 0.41 |
| Immunocompromised | 13 (0.7%) | 2 (1.5%) | 11 (0.6%) | 0.57 |
| ***Treatments and outcomes, n (%)*** |  |  |  |  |
| In-hospital deaths | 294 (16.3%) | 18 (13.4%) | 276 (16.6%) | 0.41 |
| Admitted to ICU | 331 (18.4%) | 15 (11.2%) | 316 (18.9%) | 0.034 |
| Invasive mechanical ventilation | 222 (12.3%) | 12 (8.9%) | 210 (12.6%) | 0.27 |
| Renal replacement therapy | 198 (11.0%) | 10 (7.5%) | 188 (11.3%) | 0.22 |

^1^ Mood’s test for medians or Chi-squared test for categorical data
^2^See eTable 3 for ICD-10 code lists
^3^Bisoprolol, Atenolol, Propranolol, Carvedilol.

#### eTable 2. Patient characteristics following first positive SARS-CoV-2 test, by value of first aldosterone thereafter.

| **Characteristic** | **Aldosterone <= 70 pmol/L** | **Aldosterone > 70 pmol/L** | **p-value^1^** |
| --- | --- | --- | --- |
| Sample size (*n*) | 74 | 52 |  |
| Age at admission, median [IQR] (% missing) | 64 [48.25, 82] (0%) | 58 [41, 77] (0%) | 0.28 |
| Gender (male), *n* (%) | 43 (58.1%) | 30 (57.7%) | ~1 |
| ***Ethnicity, n (%)*** |  |  |  |
| White | 46 (62.2%) | 36 (69.2%) | 0.529 |
| Black | 1 (1.4%) | 0 (0%) | ~1 |
| Asian | 9 (12.2%) | 4 (7.7%) | 0.607 |
| Other | 2 (2.7%) | 1 (1.9%) | ~1 |
| Not specified/prefer not to say | 16 (21.6%) | 11 (21.2%) | ~1 |
| Body mass index, median [IQR] (% missing), kg/m | 25.4 [23.2, 29.6] (9.5%) | 28.6 [24.75, 32.85] (9.6%) | 0.14 |
| ***Observations, median [IQR] (% missing)*** |  |  |  |
| Heart rate, beats/min | 85 [75, 96] (1.4%) | 88 [77, 95] (0%) | 0.61 |
| Temperature, ℃ | 37.4 [36.8, 38.1] (0%) | 37.1 [36.7, 37.7] (0%) | 0.75 |
| Respiratory rate, breaths/min | 19 [17, 22] (0%) | 18 [17, 20] (0%) | 0.058 |
| Oxygen saturation (SpO2), % | 95 [94, 97] (1.4%) | 96 [94, 97] (0%) | 0.40 |
| Mean arterial pressure, mmHg | 90 [80, 100] (0%) | 91 [83, 97] (0%) | 0.11 |
| ***Blood tests, median [IQR] (% missing)*** |  |  |  |
| C-reactive protein, mg/L | 54 [26, 114] (2.7%) | 39 [25, 77] (5.8%) | 0.45 |
| White cell count, 10^9^/L | 5.9 [4.8, 6.8] (2.7%) | 5.3 [4.4, 7.1] (1.9%) | 0.85 |
| Sodium, mmol/L | 136 [134, 139] (4.1%) | 137 [134, 139] (5.8%) | 0.89 |
| Potassium, mmol/L | 4.0 [3.8, 4.3] (1.5%) | 4.1 [3.7, 4.6] (1.5%) | 0.13 |
| Neutrophils, 10^9^/L | 4.3 [3.2, 5.4] (2.7%) | 3.8 [2.6, 5.5] (1.9%) | 0.34 |
| Lymphocytes, 10^9^/L | 0.84 [0.55, 1.12] (2.7%) | 1.07 [0.71, 1.61] (1.9%) | 0.44 |
| Interleukin-6, pg/ml | 9.5 [5.73, 20.89] (59.5%) | 7.15 [3.86, 13.16] (50%) | 0.40 |
| Urea, mmol/L | 5.4 [3.7, 8.5] (10.8%) | 5.9 [4.4, 7.3] (15.4%) | 0.56 |
| Creatinine, 𝜇mol/L | 72 [59, 89] (4.1%) | 76 [64, 88] (3.8%) | 0.92 |
| D-Dimer, ng/ml | 233 [176, 373] (41.9%) | 205 [155, 362] (50%) | 0.37 |
| Troponin, ng/L | 9 [3.1, 32.9] (41.9%) | 4 [3, 7.5] (40.4%) | 0.95 |
| pH value | 7.41 [7.38, 7.44] (39.2%) | 7.4 [7.37, 7.44] (30.8%) | 0.94 |
| *Medical history^2^, n (%)* |  |  |  |
| Heart disease | 12 (16.2%) | 9 (17.3%) | ~1 |
| Hypertension | 23 (31.1%) | 12 (23.1%) | 0.43 |
| Beta blockers at admission^3^ | 6 (8.1%) | 3 (5.75%) | 0.88 |
| Diabetes | 19 (25.7%) | 10 (19.2%) | 0.53 |
| Endocrine disease (other than diabetes) | 7 (9.5%) | 6 (11.5%) | 0.94 |
| Stroke | 2 (2.7%) | 1 (1.9%) | ~1 |
| Dementia | 5 (6.8%) | 2 (3.8%) | 0.76 |
| Asthma | 6 (8.1%) | 8 (15.4%) | 0.32 |
| Respiratory disease (other than asthma) | 8 (10.8%) | 5 (9.6%) | ~1 |
| Chronic kidney disease | 4 (5.4%) | 2 (3.8%) | ~1 |
| Chronic liver disease | 4 (5.4%) | 4 (7.7%) | 0.88 |
| Malignancy non-haematological | 13 (17.6%) | 3 (5.8%) | 0.092 |
| Malignancy haematological | 4 (5.4%) | 0 (0%) | 0.24 |
| Immunocompromised | 2 (2.7%) | 0 (0%) | 0.64 |
| ***Treatments and outcomes, n (%)*** |  |  |  |
| In-hospital deaths | 10 (13.5%) | 6 (11.5%) | 0.96 |
| Admitted to ICU | 9 (12.2%) | 7 (13.5%) | 0.96 |
| Invasive mechanical ventilation | 7 (9.5%) | 7 (13.5%) | 0.68 |
| Renal replacement therapy | 6 (8.1%) | 4 (7.7%) | ~1 |

^1^Mood’s test for median values, Chi-squared test for categorical data

^2^See eTable 3 for ICD-10 code lists
^3^Bisoprolol, Atenolol, Propranolol, Carvedilol.

#### eTable 3. International Classification of Diseases 10th edition (ICD-10) codes used to identify comorbidities.

| **Diagnosis** | **ICD-10 codes** | **Description** |
| --- | --- | --- |
| **Heart disease** | I20 | Angina pectoris |
|  | I21 | Acute myocardial infarction |
|  | I22 | Subsequent myocardial infarction |
|  | I23 | Certain current complications following acute myocardial infarction |
|  | I24 | Other acute ischaemic heart diseases |
|  | I25 | Chronic ischaemic heart disease |
|  | I34 | Nonrheumatic mitral valve disorders |
|  | I35 | Nonrheumatic aortic valve disorders |
|  | I36 | Nonrheumatic tricuspid valve disorders |
|  | I37 | Pulmonary valve disorders |
|  | I42 | Cardiomyopathy |
|  | I43 | Cardiomyopathy in diseases classified elsewhere |
|  | I44 | Atrioventricular and left bundle-branch block |
|  | I50 | Heart failure |
| **Hypertension** | I10 | Essential hypertension |
|  | I11 | Hypertensive heart disease |
|  | I12 | Hypertensive renal disease |
|  | I13 | Hypertensive heart and renal disease |
|  | I15 | Secondary hypertension |
| **Diabetes** | E10 | Type 1 diabetes mellitus |
|  | E11 | Type 2 diabetes mellitus |
|  | E12 | Malnutrition-related diabetes mellitus |
|  | E13 | Other specified diabetes mellitus |
|  | E14 | Other unspecified diabetes mellitus |
| **Endocrine disease (other than diabetes)** | E00 | Congenital iodine-deficiency syndrome |
|  | E01 | Iodine-deficiency-related thyroid disorders and allied conditions |
|  | E02 | Subclinical iodine-deficiency hypothyroidism |
|  | E03 | Other hypothyroidism |
|  | E04 | Other nontoxic goitre |
|  | E05 | Thyrotoxicosis |
|  | E06 | Thyroiditis |
|  | E07 | Other disorders of thyroid |
|  | E20 | Hypoparathyroidism |
|  | E21 | Hyperparathyroidism and other disorders of parathyroid gland |
|  | E22 | Hyperfunction of pituitary gland |
|  | E23 | Hypofunction and other disorders of pituitary gland |
|  | E24 | Cushing syndrome |
|  | E25 | Adrenogenital disorders |
|  | E26 | Hyperaldosteronism |
|  | E27 | Other disorders of adrenal gland |
|  | E28 | Ovarian dysfunction |
|  | E29 | Testicular dysfunction |
|  | E30 | Disorders of puberty, not elsewhere classified |
|  | E31 | Polyglandular dysfunction |
|  | E32 | Diseases of thymus |
|  | E34 | Other endocrine disorders |
|  | E35 | [Disorders of endocrine glands in diseases classified elsewhere](https://icd.codes/icd10cm/chapter4/E20-E35) |
|  | E89.0 | [Postprocedural hypothyroidism](https://icd.codes/icd10cm/E890) |
|  | E89.1 | [Postprocedural hypoinsulinemia](https://icd.codes/icd10cm/E891) |
|  | E89.2 | [Postprocedural hypoparathyroidism](https://icd.codes/icd10cm/E892) |
|  | E89.3 | [Postprocedural hypopituitarism](https://icd.codes/icd10cm/E893) |
|  | E89.4 | [Postprocedural ovarian failure](https://icd.codes/icd10cm/E894) |
|  | E89.5 | [Postprocedural testicular hypofunction](https://icd.codes/icd10cm/E895) |
|  | E89.6 | [Postprocedural adrenocortical (-medullary) hypofunction](https://icd.codes/icd10cm/E896) |
| **Stroke** | I63 | Cerebral infarction |
|  | I65 | Occlusion and stenosis of precerebral arteries, not resulting in cerebral infarction |
|  | I66 | Occlusion and stenosis of cerebral arteries, not resulting in cerebral infarction |
| **Dementia** | F01 | Vascular dementia |
|  | F02 | Dementia in other diseases classified elsewhere |
|  | F03 | Unspecified dementia |
|  | G30, G31 | Alzheimer disease & Other degenerative diseases of nervous system, not elsewhere classified |
|  | F10.27 | Alcohol dependence, with alcohol-induced persisting dementia |
|  | F10.97 | Alcohol use, unspecified with alcohol-induced persisting dementia |
|  | F19.97 | Other psychoactive substance use, unspecified with psychoactive substance-induced persisting dementia |
| **Asthma** | J45 | Asthma |
| **Respiratory disease (other than asthma)** | I27 | Other pulmonary heart diseases |
|  | J6*-J7* | Lung diseases due to external agents |
|  | J41 | Simple and mucopurulent chronic bronchitis |
|  | J42 | Unspecified chronic bronchitis |
|  | J43 | Emphysema |
|  | J44 | Other chronic obstructive pulmonary disease |
|  | J47 | Bronchiectasis |
| **Chronic kidney disease** | N18.1-N18.5 | Chronic kidney disease stage 1-5 |
|  | N18.9 | Chronic kidney disease, unspecified |
|  | I13 | Hypertensive and renal disease |
| **Chronic liver disease** | K70 | Alcoholic liver disease |
|  | K71 | Toxic liver disease |
|  | K72 | Hepatic failure, not elsewhere classified |
|  | K73 | Chronic hepatitis, not elsewhere classified |
|  | K74 | Fibrosis and cirrhosis of the liver |
|  | K75 | Other inflammatory diseases of the liver |
|  | K76 | Other diseases of the liver |
|  | K77 | Liver disorders in disease classified elsewhere |
| **Malignancy non-haematological** | C0 | Malignant neoplasm of lip |
|  | C1 | Malignant neoplasm of base of tongue |
|  | C2 | Malignant neoplasm of other unspecified parts of tongue |
|  | C3 | Malignant neoplasm of gum |
|  | C4 | Malignant neoplasm of floor of mouth |
|  | C5 | Malignant neoplasm of palate |
|  | C6 | Malignant neoplasm of other and unspecified parts of mouth |
|  | C7 | Malignant neoplasm of parotid gland |
| **Malignancy haematological** | C8 | Malignant neoplasm of other and unspecified major salivary glands |
|  | C9 | Malignant neoplasm of tonsil |
| **Immunocompromised** | D80 | Immunodeficiency with predominantly antibody defects |
|  | D81 | Combined immunodeficiencies |
|  | D82 | Immunodeficiency associated with other major defects |
|  | D83 | Common variable immunodeficiency |
|  | D84 | Other immunodeficiencies |

#### eFigure 1. Scatter plots of paired aldosterone and renin results with linear regression line, with the two extreme outliers shown included.

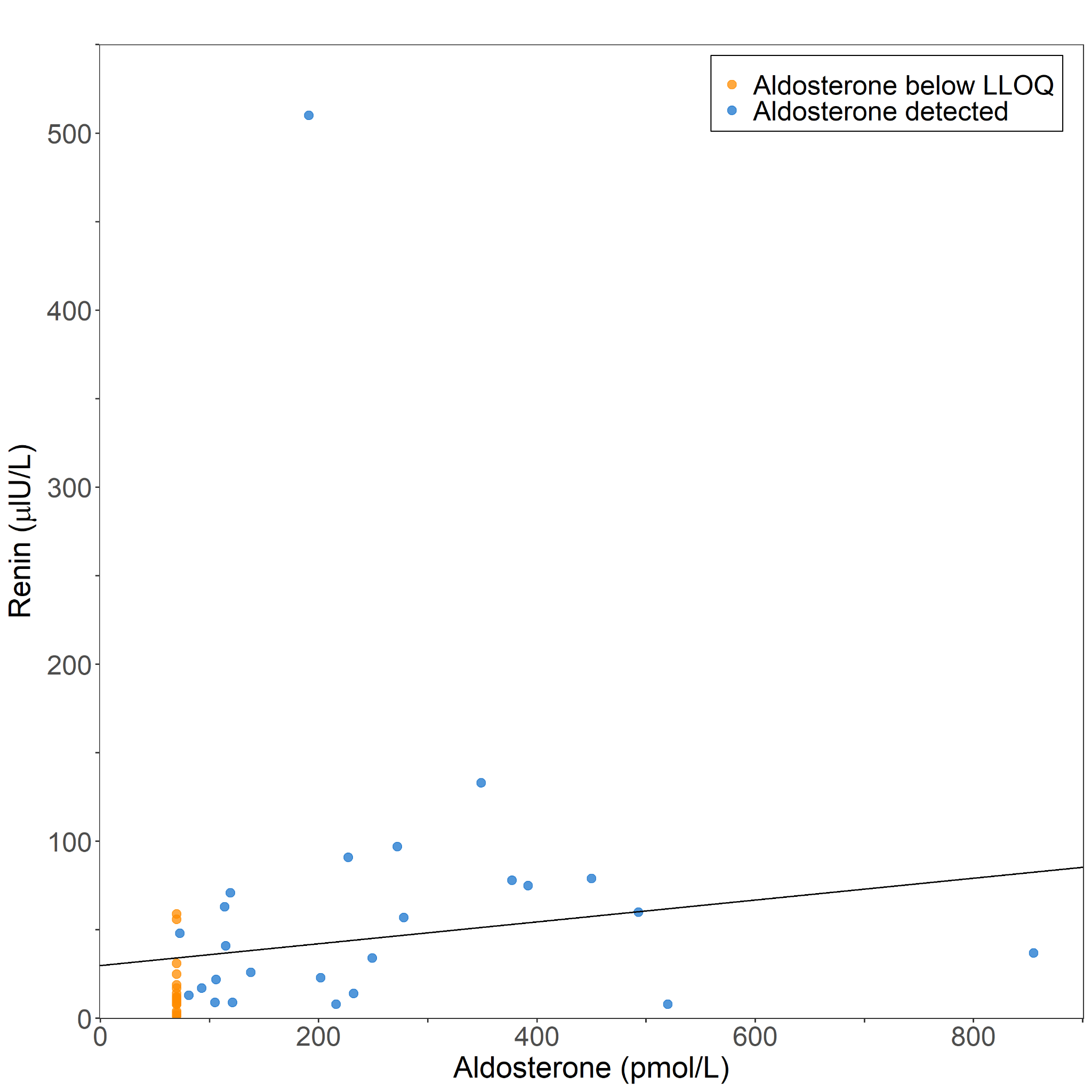

#### eFigure 2. Kaplan-Meier survival plot for 28-day survival after first positive test, stratified by whether the first aldosterone after the positive test was above or below the LLOQ.

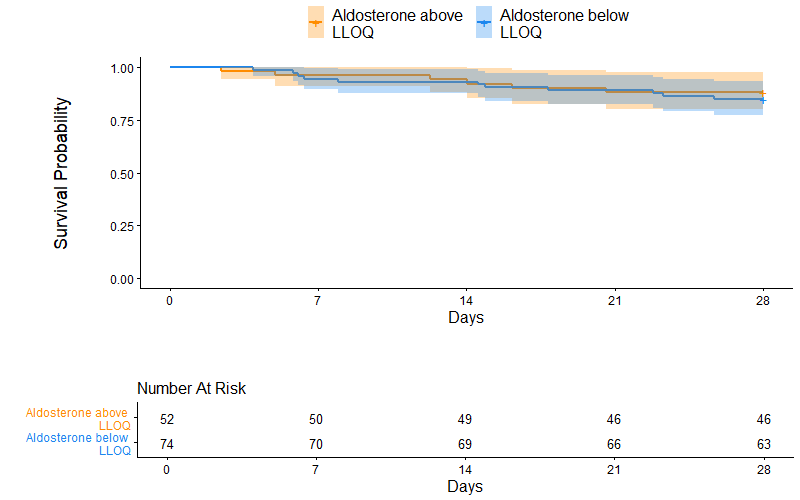

#### eFigure 3. Association plots of the LCMSMS aldosterone measurements (y-axis) and (A) eGFR (B) creatinine clearance (Cockcroft-Gault). The solid lines indicate (robust) linear regression.

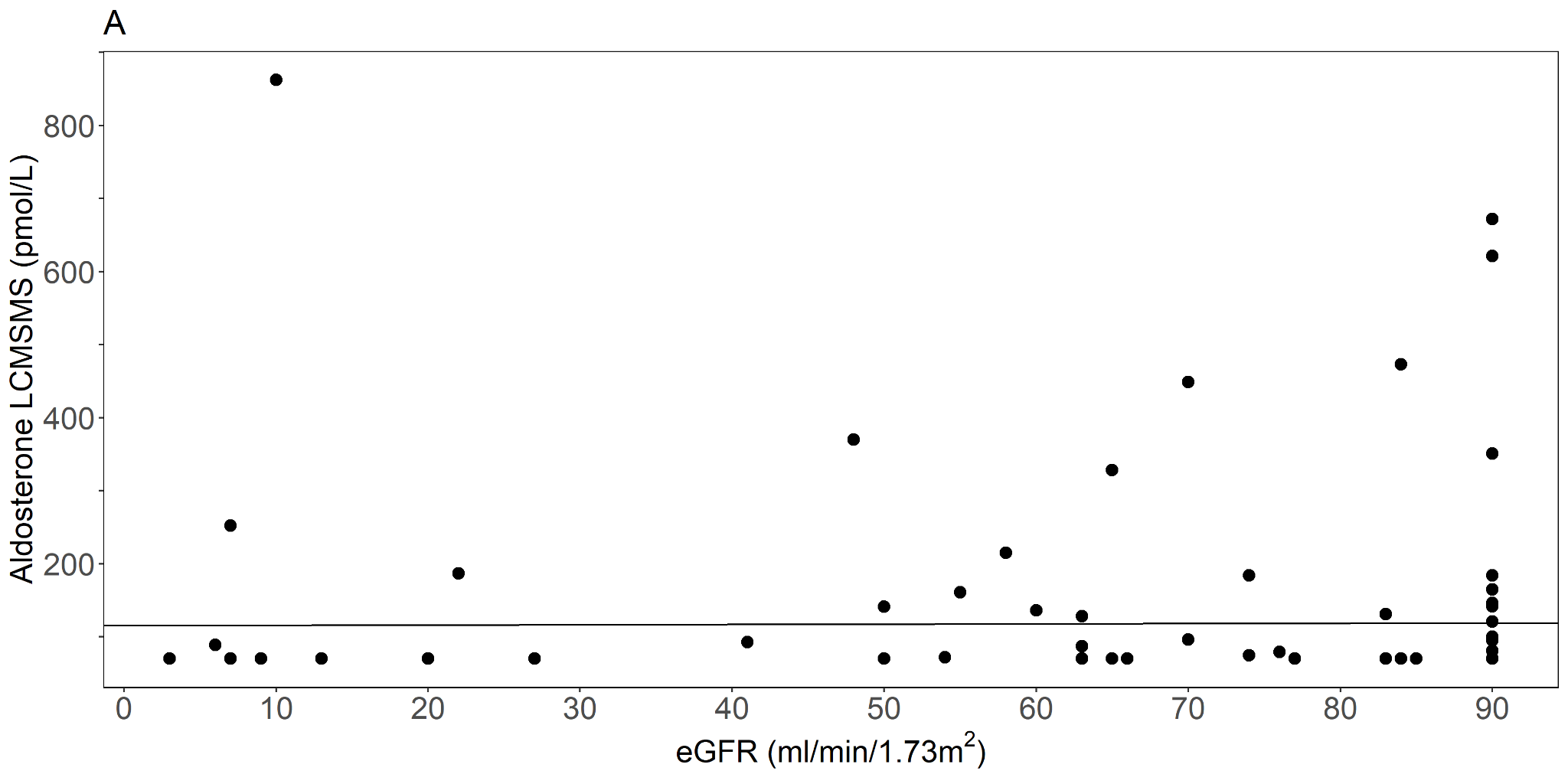

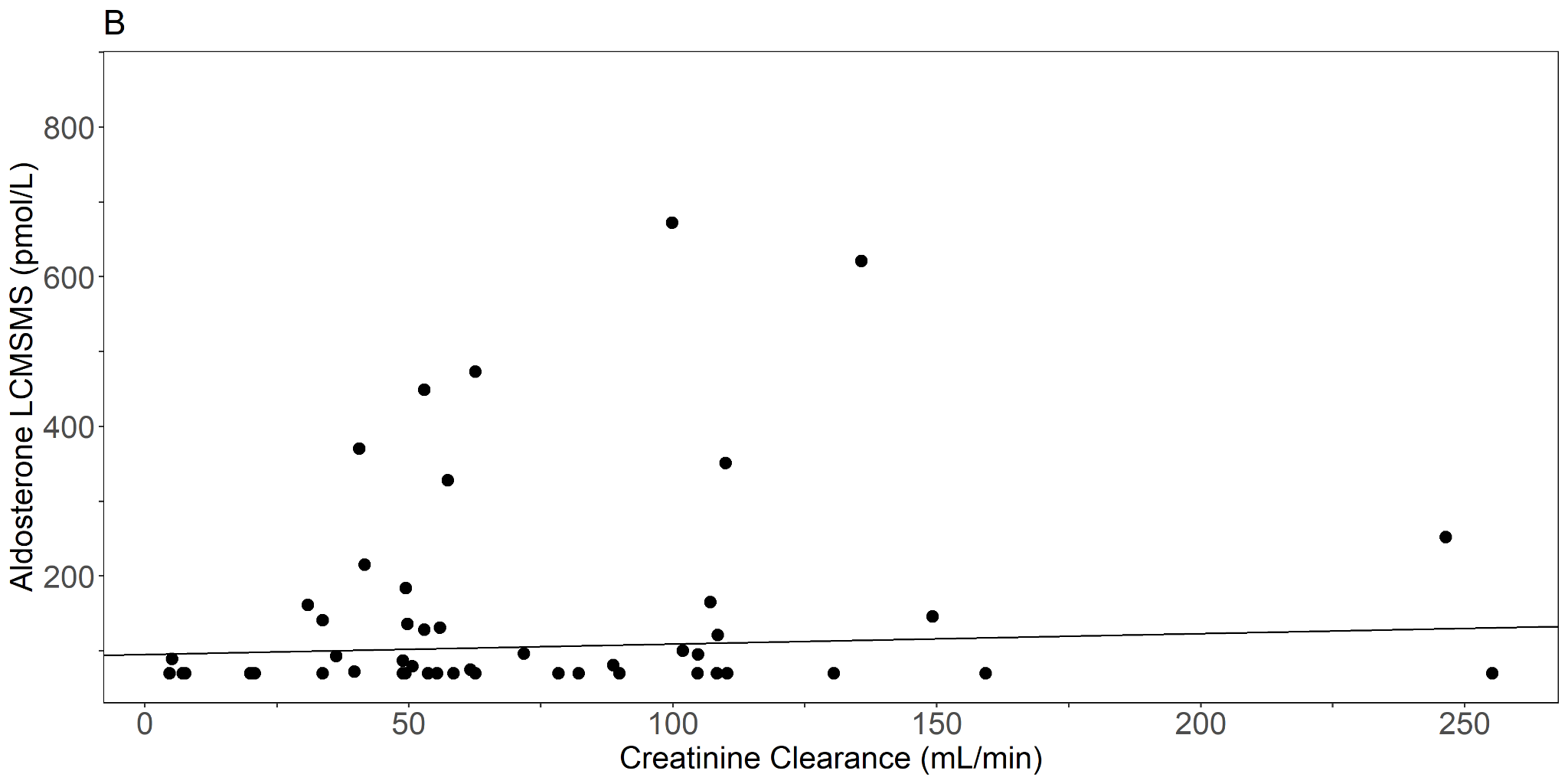

#### eFigure 4. Bland-Altman plot showing the average of the aldosterone concentration measured by CLIA and LCMSMS on the x-axis and the raw difference between methods on the y-axis.

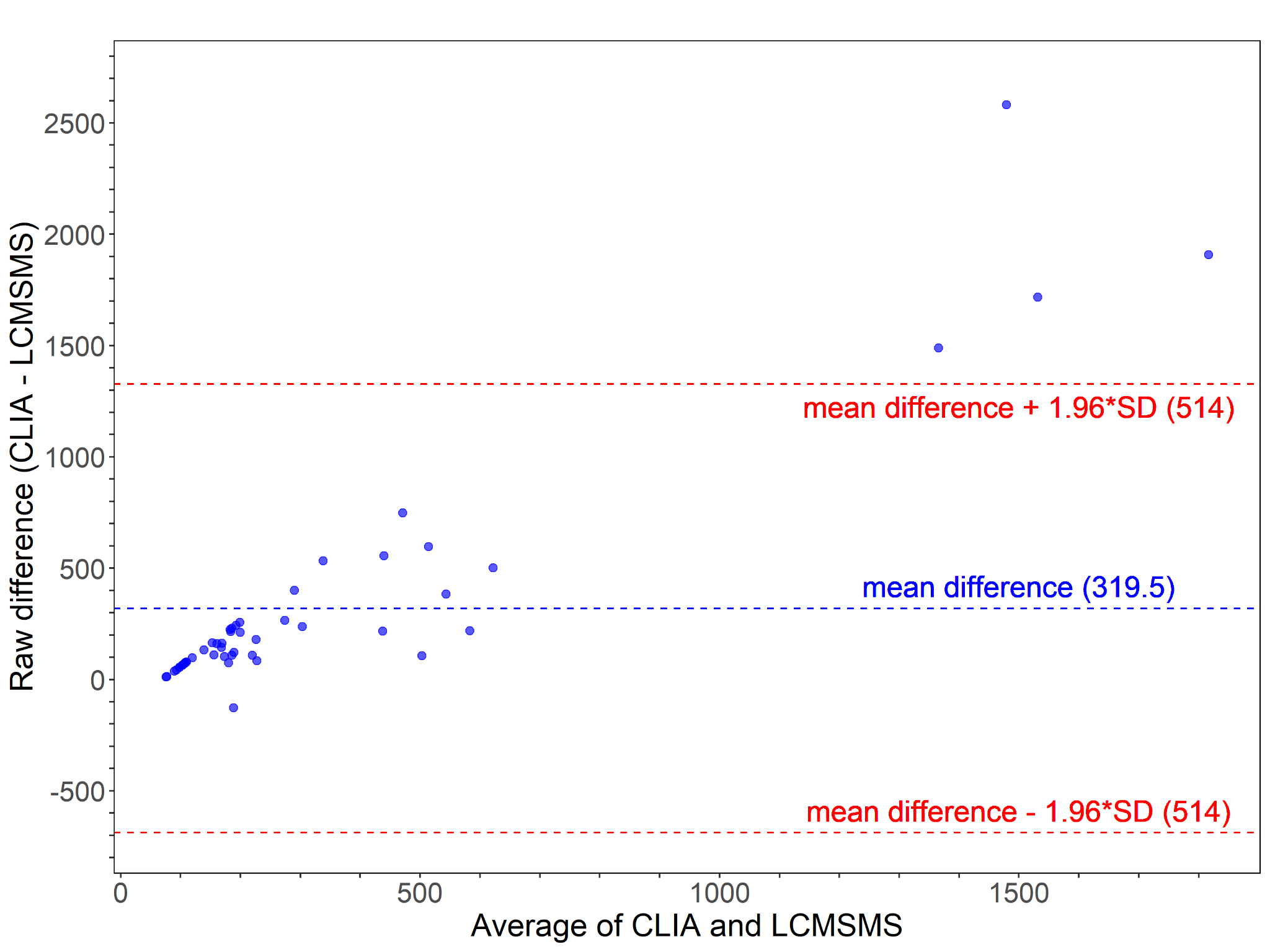

#### eFigure 5. Association plots of the absolute difference between LCMSMS and CLIA aldosterone measurements (y-axis) and (A) eGFR (B) creatinine clearance (Cockcroft-Gault) (C) bilirubin (D) ALT and (E) Cortisol. The solid lines indicate (robust) linear regression.

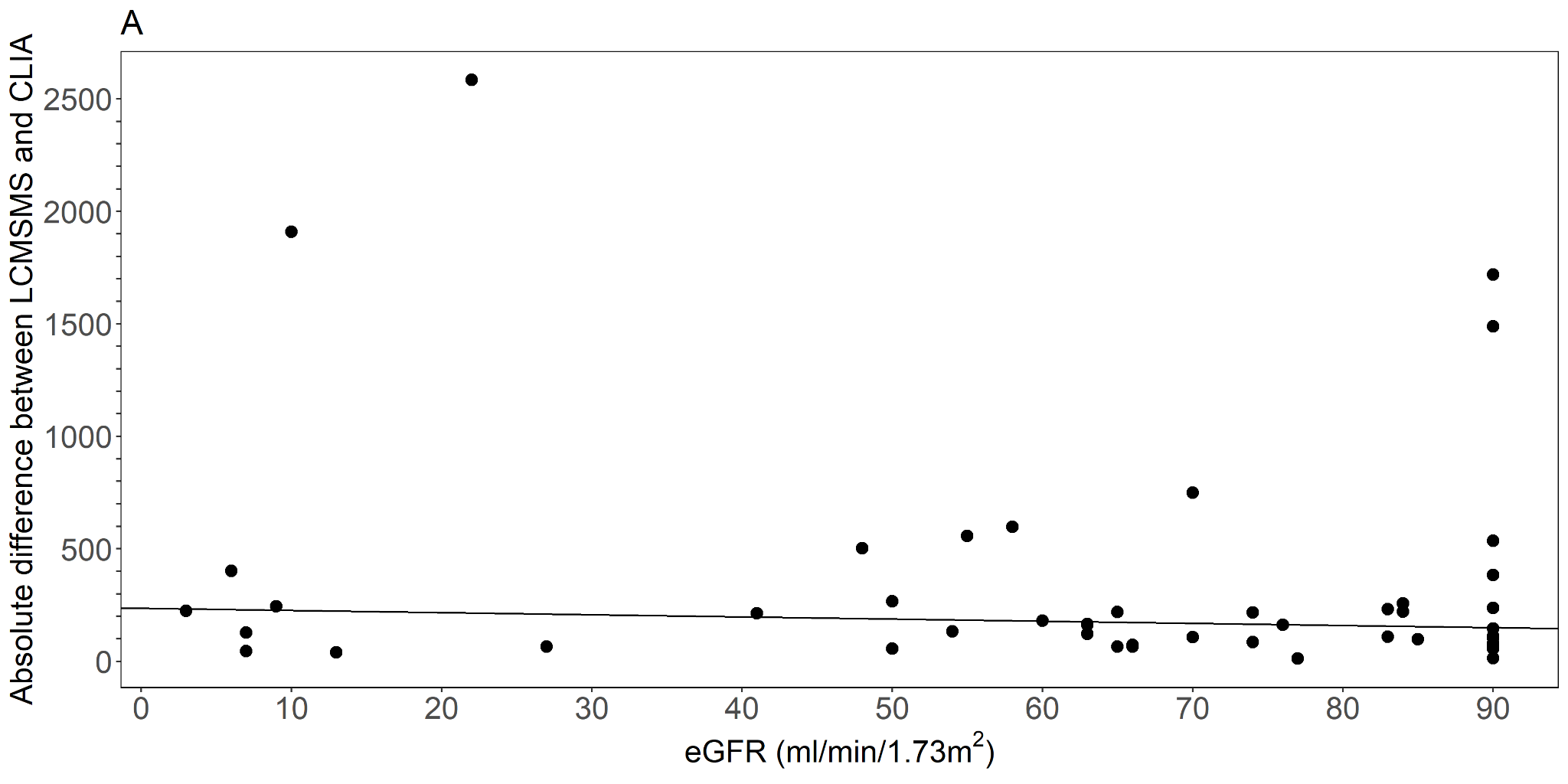

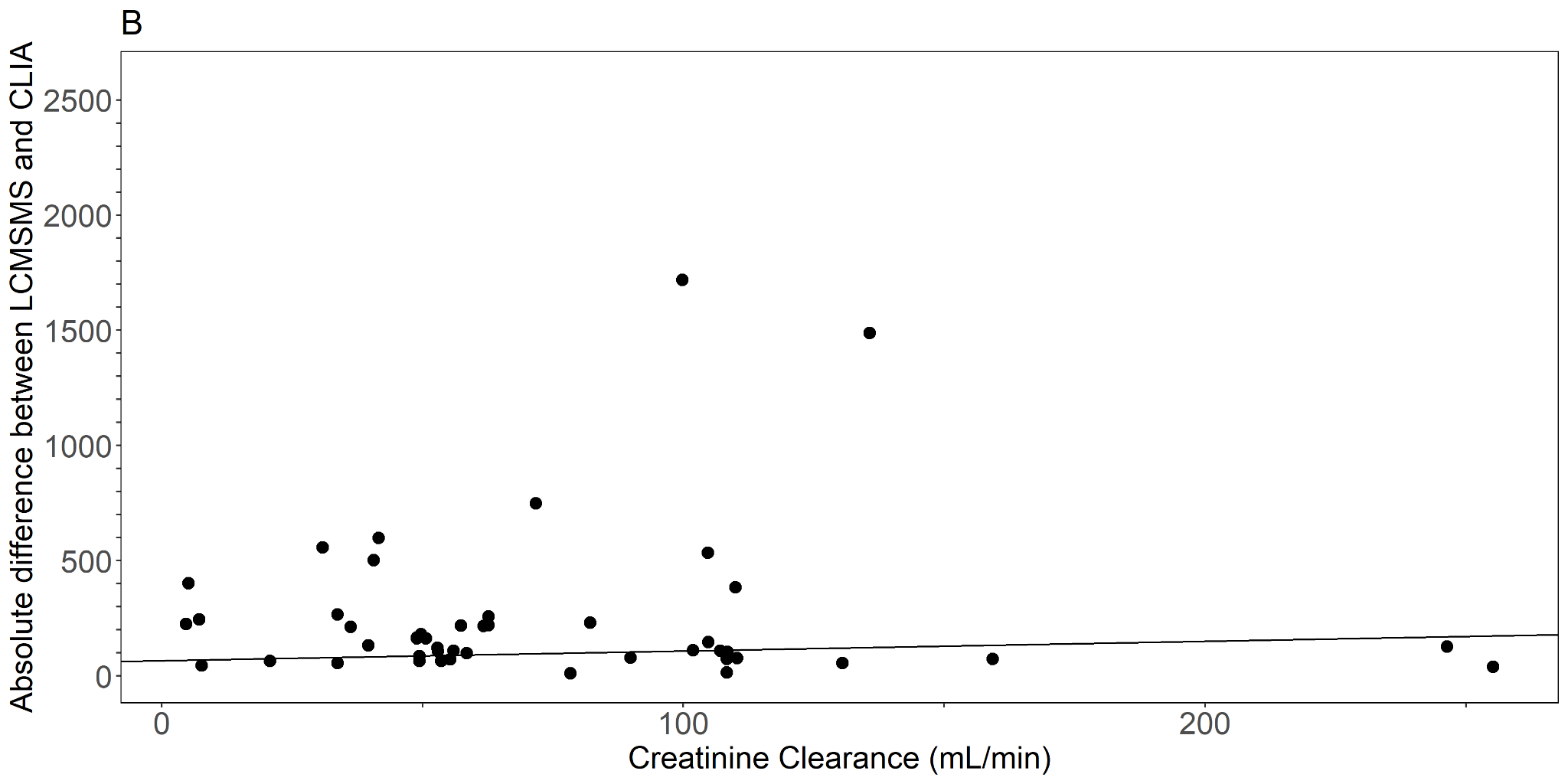

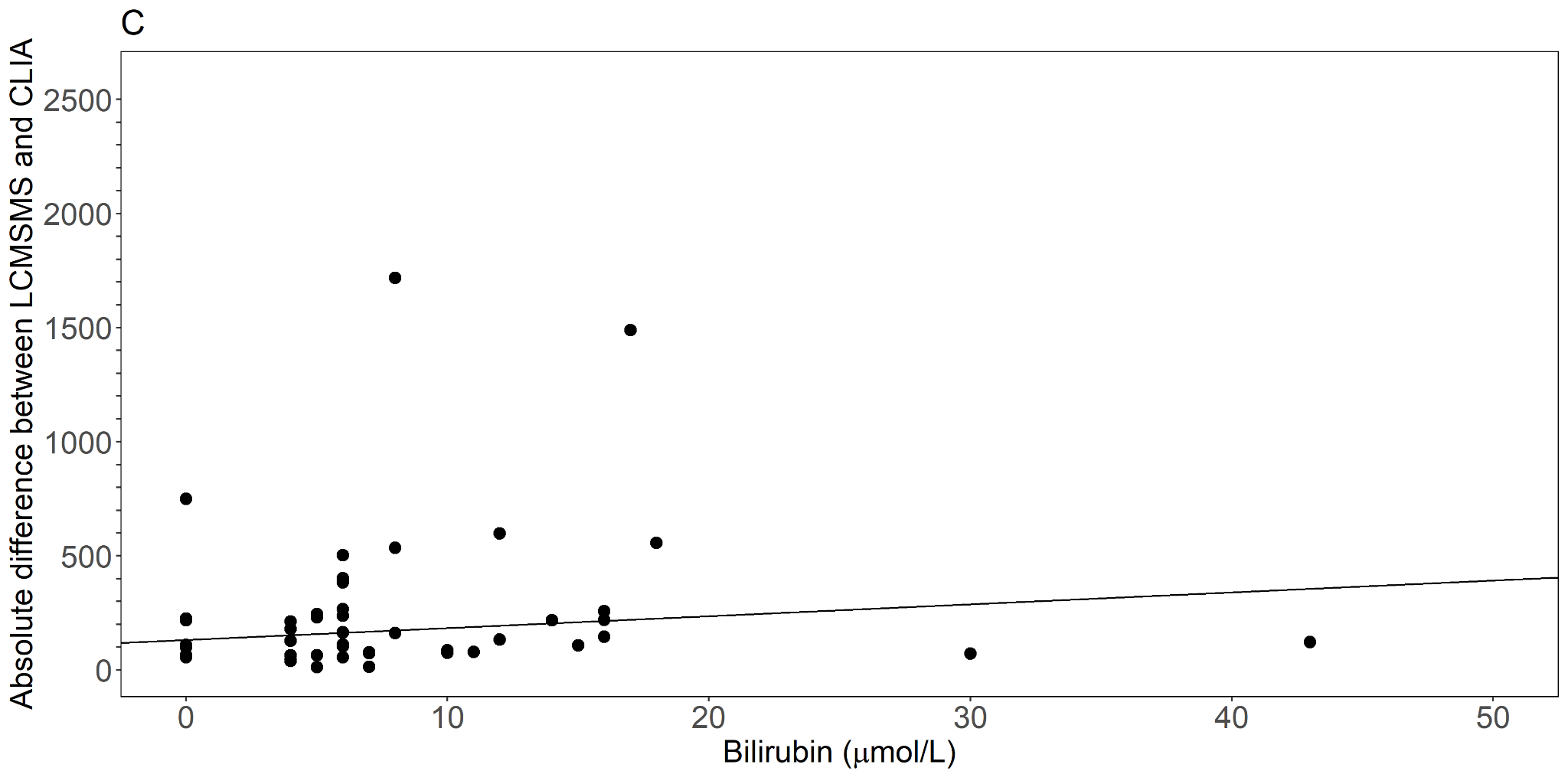

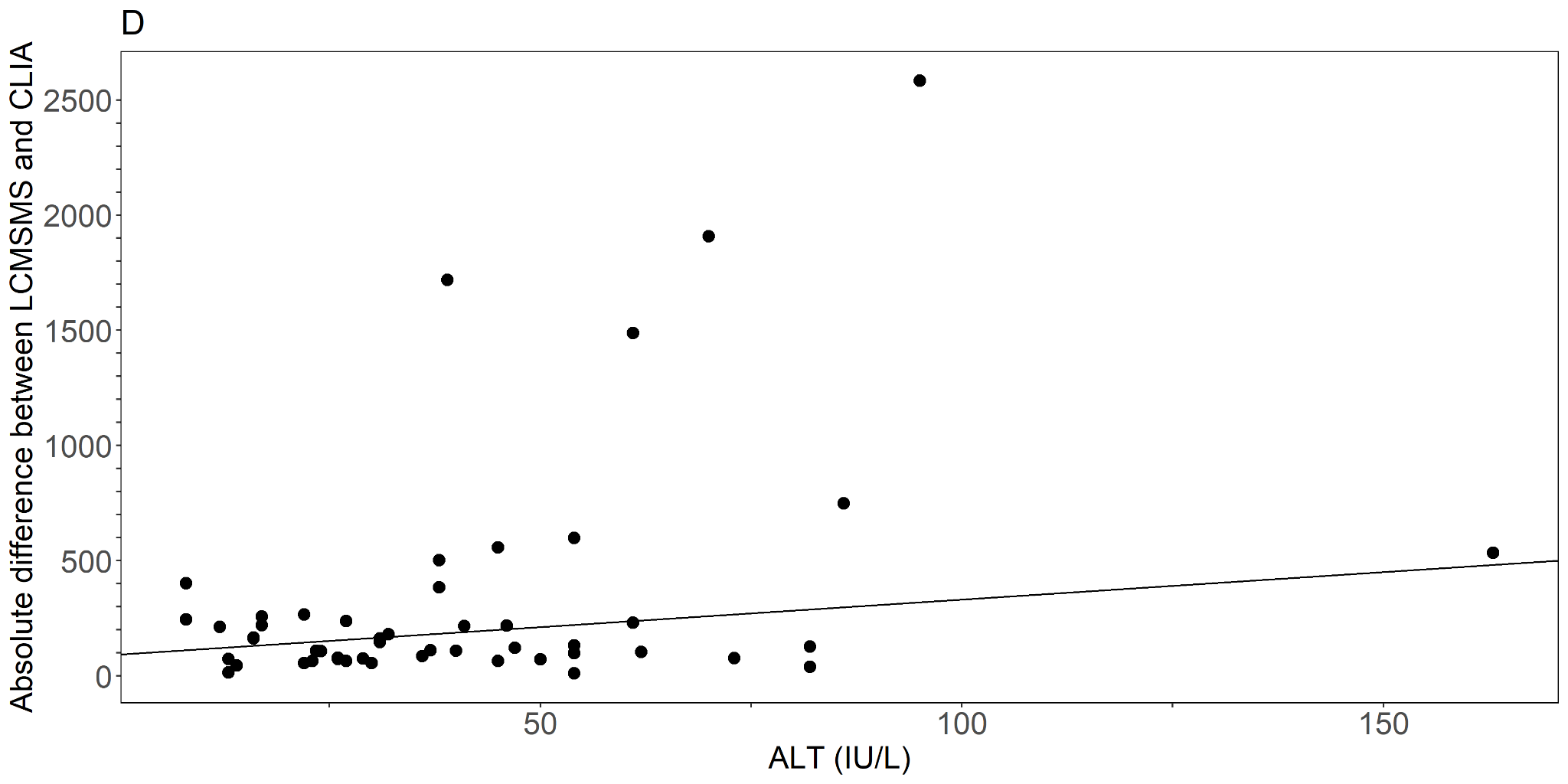

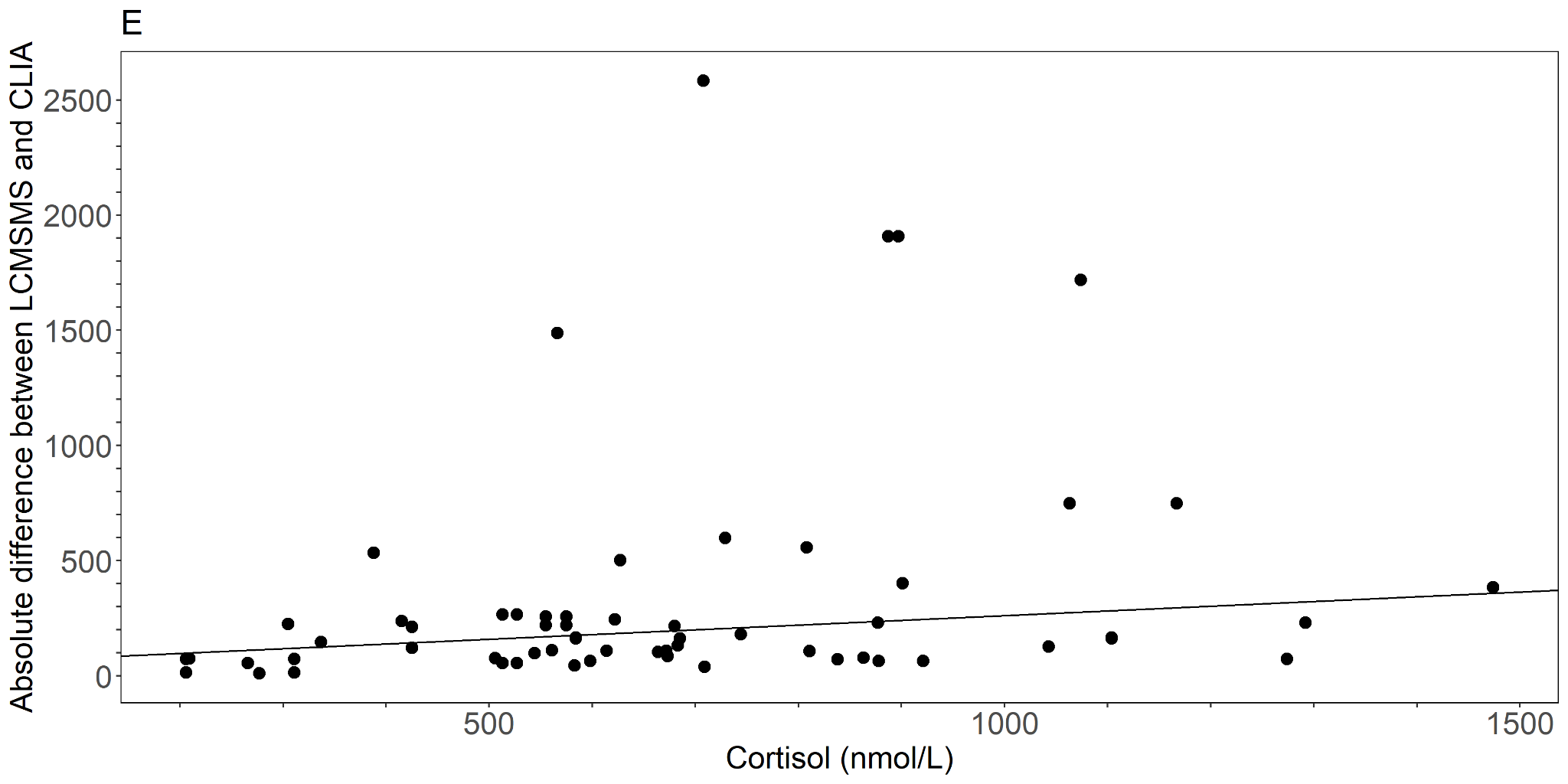

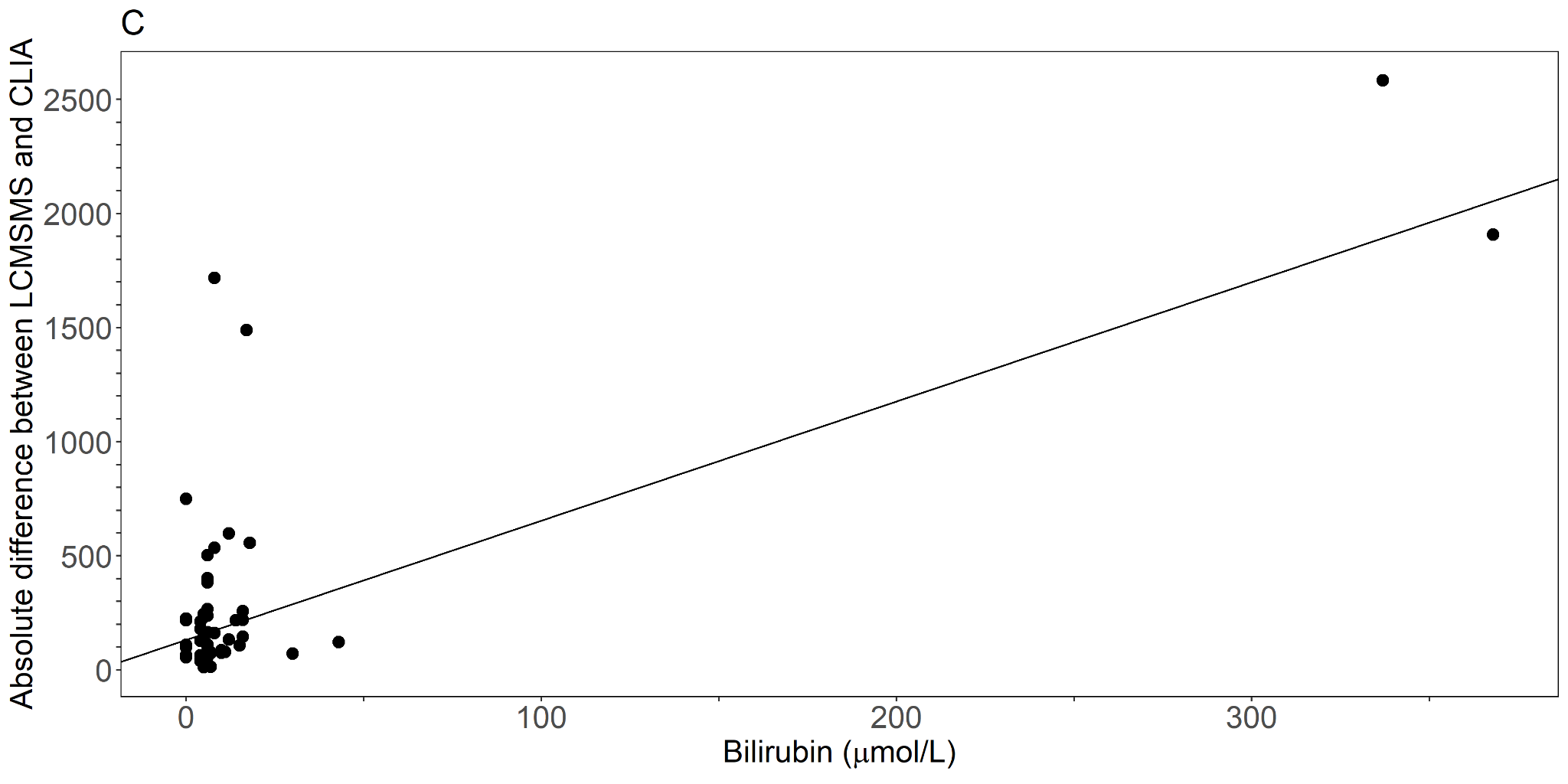

#### eFigure 6. Comparison of LCMSMS (x-axis) and CLIA aldosterone (y-axis) results, with solid lines indicating Passing-Bablok regression lines, solvent extracted (red) and non-extracted (gray). The dotted line indicates the y=x identity line.

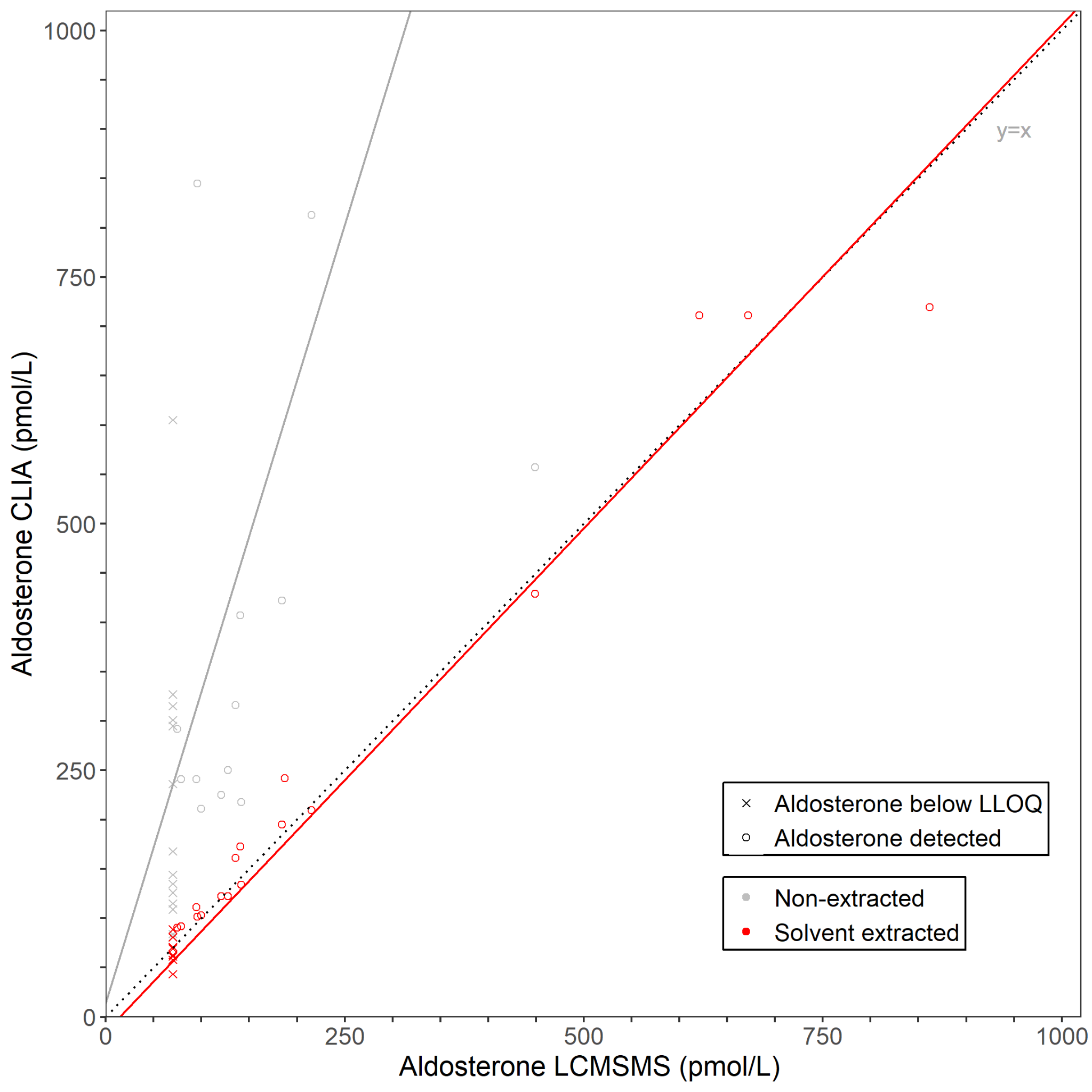
